# Safety-Critical Control of Active Interventions for COVID-19 Mitigation

**DOI:** 10.1101/2020.06.17.20133264

**Authors:** Aaron D. Ames, Tamás G. Molnár, Andrew W. Singletary, Gábor Orosz

## Abstract

The world has recently undergone the most ambitious mitigation effort in a century^1^, consisting of wide-spread quarantines aimed at preventing the spread of COVID-19^2^. The use of influential epidemiological models^3–6^ of COVID-19 helped to encourage decision makers to take drastic non-pharmaceutical interventions. Yet, inherent in these models are often assumptions that the active interventions are static, e.g., that social distancing is enforced until infections are minimized, which can lead to inaccurate predictions that are ever evolving as new data is assimilated. We present a methodology to dynamically guide the active intervention by shifting the focus from viewing epidemiological models as systems that evolve in autonomous fashion to control systems with an “input” that can be varied in time in order to change the evolution of the system. We show that a safety-critical control approach^7^ to COVID-19 mitigation gives active intervention policies that formally guarantee the safe evolution of compartmental epidemiological models. This perspective is applied to current US data on cases while taking into account reduction of mobility, and we find that it accurately describes the current trends when time delays^8^ associated with incubation and testing are incorporated. Optimal active intervention policies are synthesized to determine future mitigations necessary to bound infections, hospitalizations, and death, both at national and state levels. We therefore provide means in which to model and modulate active interventions with a view toward the phased reopenings that are currently beginning across the US and the world in a decentralized fashion. This framework can be converted into public policies, accounting for the fractured landscape of COVID-19 mitigation in a safety-critical fashion.

## Introduction

As COVID-19 spreads throughout the world^9–11^, due to the novelty of the virus and the resulting lack of pharmaceutical options necessary to suppress infection^12^, unprecedented mitigation steps to slow its progression were taken in the form of non-pharmaceutical interventions^3,13^, e.g., social distancing, mask-wearing, quarantining, and stay-at-home orders. It is largely agreed upon that these slowed the spread of the virus^2,14^, thereby saving lives. Yet studies have shown that if these active interventions had been enforced even a week earlier^15^, the result would have been a substantial reduction in deaths. As a means of mitigating the spread of COVID-19, the question therefore becomes: when, where, and how does one decide to take non-pharmaceutical interventions? This question is especially relevant^16^ as restrictions are being relaxed in a decentralized fashion across the US and throughout the world.

Due to the pressing need to understand past and future mitigation efforts, and the corresponding role of active interventions, there has been a surge of recent papers on the modeling of COVID-19^5,17–20^. Epidemiological models for predicting the spread of COVID-19 often utilize dynamical systems obtained from so-called “compartmental” models wherein the compartments are chosen to reflect different populations of interest^21–23^, e.g., susceptible (*S*), infected (*I*), recovered (*R*), etc. More compartments can be added allowing for higher fidelity models, although one must be careful of overfitting the largely increased number of parameters in more complex models. The most fundamental (and elementary) of these compartmental models is the SIR model, which has recently been used in modeling of COVID-19^24^. Examples of more complex compartmental models applied for COVID-19 include the SEIR^25, 26^ and SIRT^27^ models, which involve exposed (*E*) and threatened (*T*) populations, and the SIDARTHE model^5^ which adds even more compartments. While these models have been found to to be useful when modeling the spread of COVID-19 and the corresponding mitigation procedures, e.g., stay-at-home orders, the approaches are fundamentally based on autonomous dynamics^28,29^ as they do not have a time-varying *control input* that can dynamically change the evolution of the system. We propose a different approach: applying *safety-critical control* methods to guide active non-pharmaceutical interventions wherein we can actively predict the interventions needed to maintain safety by viewing compartmental models as *control systems*.

### SIR model as a Control System

At the core of our approach is a fundamental shift in perspective on epidemiological models: from viewing them as dynamical systems that evolve in an autonomous fashion, to that of control systems for which the evolution can be dynamically modified. In many ways, this is the *de facto* manner in which these models are implemented, if only in an implicit fashion, as they are constantly updated as new data is assimilated, e.g., as changes in social distancing are observed, predictive models are updated^30^. We, therefore, will formalize this perspective by making the control aspect of epidemiological models explicit. Note that viewing compartmental epidemiological models as control systems is not unique^31,32^, but has found only limited application to COVID-19^33^ and has yet to enjoy formal guarantees on safety. Additionally, there are examples of control-theoretic concepts being applied, namely in the the context of time-varying^27,34^ and state-varying^6,35^ choices of the transmission rate; these can be viewed as time- and state-varying inputs to a control system. Our approach differs in that we wish to synthesize *active intervention policies* (i.e., feedback control laws) that will determine future actions to take based upon past observations of the states of the systems.

To motivate the methodology utilized throughout this paper, we will begin by considering the fundamental epidemiological compartmental model: the SIR model ^21,23^. Importantly, the approach introduced herein can be applied to *any* compartmental model, and will subsequently be applied to a more descriptive model. The SIR model consists of a *susceptible* population *S, infected* population *I*, and *recovered* population *R*. We can view the evolution of these populations as a control system where active interventions, expressed by the control input *u*(*t*), modulate the rate of change of the infected population:

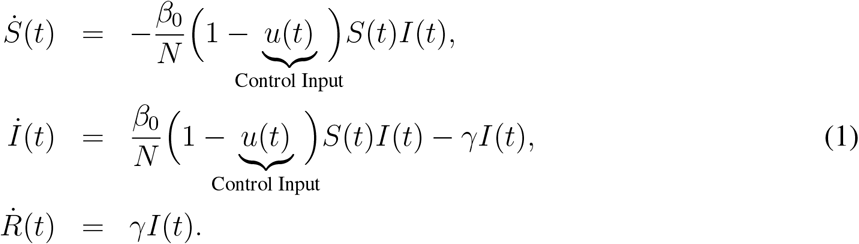

Here the total population *N* = *S*(*t*)+*I*(*t*)+*R*(*t*) is constant, *β*_0_ > 0 is the transmission rate (when no intervention is present) and *γ* > 0 is the recovery rate, yielding the reproduction number: *R*_0_ = *β*_0_*/γ*. This model relates to the traditional SIR model via the time-varying transmission rate *β*(*t*) = *β*_0_(1 − *u*(*t*)). Time-varying *β*(*t*) has been considered^20^; for example, we can utilize the policy *u*(*t*) = −*A* cos(*ωt*) in the SIR model^36^ to recover models of seasonal variations in infection^37^. In the setting considered here, taking *u*(*t*) ≡ 0 corresponds to *no* intervention, yielding the traditional SIR model with *β*(*t*) ≡ *β*_0_, whereas *u*(*t*) ≡ 1 can be viewed as *maximum* intervention, full and complete quarantine of the population. In the latter case the infected population decays to zero exponentially, *I*(*t*) = e^−*γt*^*I*(0), since the susceptible population is isolated. These effects can be seen, for example, in the Chinese response to COVID-19 and the corresponding drop in *R*_0_ documented^19^.

An illustration of the SIR model as a control system is shown in Fig. 1 where the interactions between the compartments are denoted by arrows with appropriate rate constants indicated. The blue arrow represents the time dependent modulation of the transmission rate *β*(*t*). The control input *u*(*t*) is estimated from mobility data^38^ in the US between March 2 and May 20, 2020 by assuming *u* = 0 at the beginning this period when no non-pharmaceutical interventions were present. Furthermore, the parameters *β*_0_, *γ* and *N* of the SIR model were fitted to the recorded number of confirmed cases: *I*(*t* −*τ*) + *R*(*t* −*τ*). Fitting for the time delay *τ*, in the corresponding transmission rate *β*(*t* −*τ*), reveals that the COVID-19 data^39^ depicted publicly^40^ are delayed by *τ* ≈10 days. This time delay originates from the incubation time of the virus (i.e., people are being infectious before being symptomatic) and the time needed for testing^33,41,42^. That is, the data corresponds the number of confirmed cases *τ* days ago while the real current number could be much higher. For example, in mid-March, when interventions were introduced in the US, *I*(*t* −*τ*) + *R*(*t* −*τ*) was reported to be in the range of a few thousand, while the real number *I*(*t*) + *R*(*t*) is estimated to be more than a hundred thousand. This delay also appears in the active intervention policies which depend on the state of the system, i.e.,*u*(*t*) = *A*(*S*(*t*−*τ),I(t*−*τ)*), and it therefore must be compensated for in order to ensure the safety of these policies. Finally, we remark that when fitting the model (1) to the aforementioned data one may obtain good fits while setting *N* in the range from 7.5 million up to 330 million (see the Methods section for additional details). Smaller values encode the fact that not everyone susceptible is necessarily exposed when the total number of infected is small relative to the total population, as well as the fact that the total number of infections is underreported^43^. In Fig. 1 we used the lowest value *N* = 7.5 million; the consequences of this choice will be discussed in the context of active interventions.

**Figure 0.**
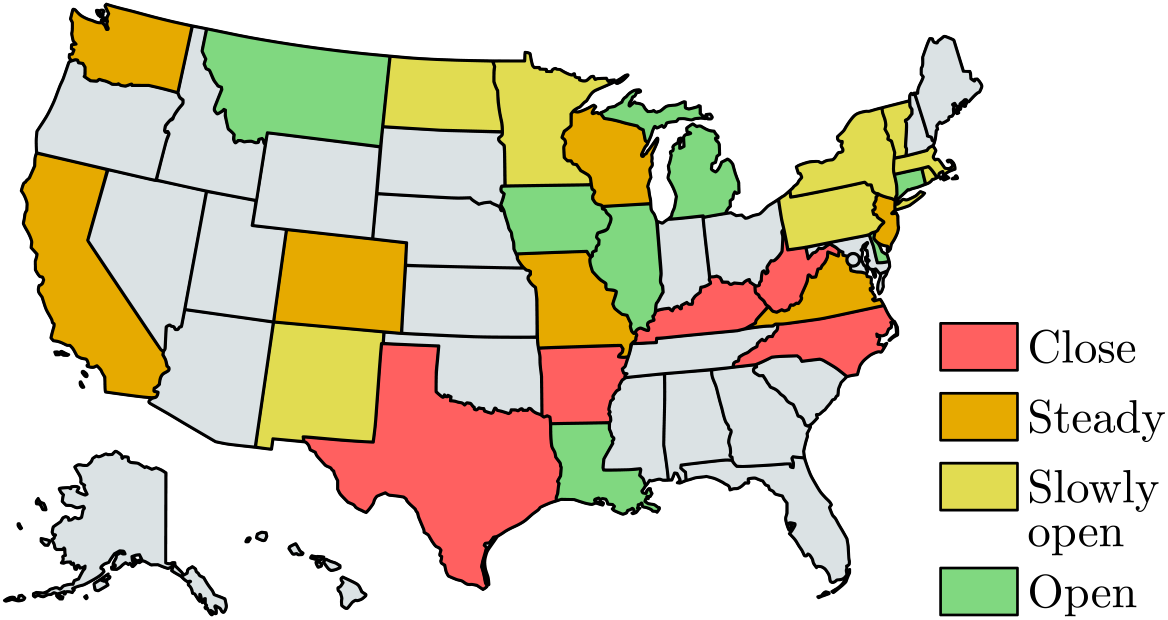
Illustration of the safety-critical active intervention policies developed in this paper applied at the state level (for states with sufficient data). The states are colored according to whether it is safe to open further (green), slowly open (yellow) hold the current mitigation efforts steady (orange), or increase mitigation (red). This is determined based upon an active intervention policy that formally guarantees bounded hospitalizations and deaths.

**Figure 1.**
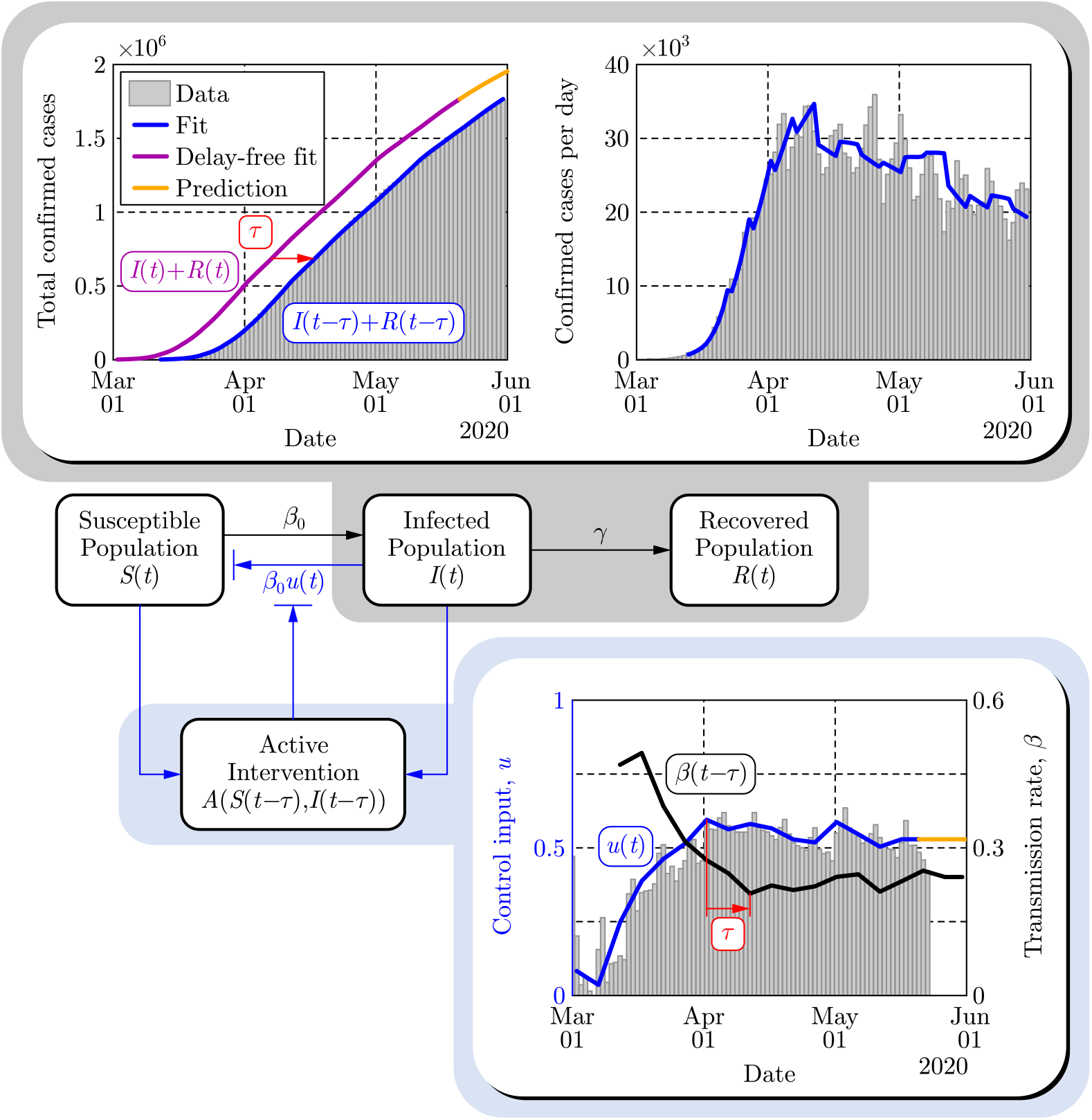
Paradigm shift wherein compartmental models are viewed as control systems rather than dynamical systems. This is illustrated on the populations *I*(*t*) and *R*(*t*) of the SIR model (top panels) wherein the control input *u*(*t*) is modulated based upon the intervention policies estimated from mobility data (bottom panel). The time delay *τ*= 10 days is highlighted to emphasize that the observed data corresponds to the delayed counterparts of the populations, and this delay also appears in the active intervention policy: *u*(*t*) = *A*(*S* (*t* − *τ*),*I*(*t* − *τ*)) given in Eq. (2).

### Safety-Critical Control for Active Intervention

Utilizing the paradigm of epidemiological models as control systems, we can synthesize active intervention policies, i.e., inputs to Eq. (1) expressed as functions of the populations of the compartmental model. A special case of this is referred to as shield immunity^6^, wherein the policy 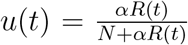 with *α ≥*0 was chosen. Our goal is to synthesize active intervention policies so as to achieve desired safety-critical behaviors, that is, to guarantee that the system, with the policy applied, evolves in a safe fashion. Concretely, we may quantify safety in the context of the SIR model as limiting the total number of infected persons: *I*(*t*) ≤ *I*_max_. To achieve such goal, we leverage the framework of control barrier functions^7^ which gives necessary and sufficient conditions on the safety, along with tools to generate active intervention policies that ensure safety.

While there may exist multiple safe policies, it is beneficial to chose one which minimizes the active intervention *u*(*t*), since more aggressive interventions potentially result in the lose of jobs and other economic and physiological effects^44, 45^. The active intervention policy, i.e., feedback control law, that gives the minimal possible (pointwise optimal) interventions so as to ensure the safety of the system can be explicitly calculated (as described in Methods):

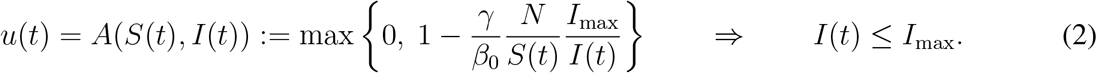

Notice the *activation function* or *rectified linear unit*^46^ ReLU(*x*) = max{0; *x*} can be used to express the policy; notably, this also used in neural networks in the context of machine learning^47^. This highlights that interventions only become “active” when safety is in danger of being violated. However, if one simply uses the obtained feedback control law in the SIR model with time delay *τ*, i.e., substitutes *u*(*t*) = *A*(*S*(*t*−*τ),I(t*−*τ)*) into Eq. (1), safety cannot be ensured due to the delay. In order to compensate for this delay we construct predictors^48^ (as described in Methods) and use the predicted states *S*_p_(*t*) and *I*_p_(*t*) in the active intervention policy: *u*(*t*) = *A*(*S*_p_ (*t*),*I*_p_(*t*)) If the predictions are accurate, i.e., *S*_p_ (*t*) = *S* (*t*) and *I*_p_ (*t*) = *I* (*t*) then the delay-free control design can ensure safety. Such predictors play an essential role in making the active intervention policies, synthesized from control barrier functions, implementable in the presence of time delay^8^.

Figure 2 depicts the results of applying the safety-critical active intervention policy in Eq. (2) to the SIR model in Eq. (1) while compensating for the 10 days delay using predictors. The control barrier function is able to keep the infected population under *I*_max_ = 200, 000 while gradually driving the control input (active intervention) to zero, i.e., mitigation methods can eventually be removed. Notice that this opening strategy decreases the control input very slowly at the beginning followed by a faster opening toward the end. As a reference we also show the results of another opening strategy where the control input is reduced to zero linearly in time. In this case the number of infections peaks at a much higher value putting a large burden on the health system. While this figure vividly illustrates the use of safety-critical active intervention, and the benefits thereof, it also predicts that all restrictions can be lifted by mid-July. This is due to the use of the simplified SIR model that was considered to illustrate the concepts presented and, more specifically, due to the fact that the model heavily depends on the *N* (chosen to be 7.5 million when fitting the data). Selecting a larger *N* would yield a longer mitigation period: the time period where active intervention is necessary, i.e., where Eq. (2) is non-zero, can be calculated as 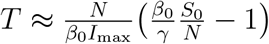, where *S*_0_ is the size of the susceptible population when the controller in Eq. (2) is initiated. Increasing *N* increases the period for which active intervention is necessary i.e., when the safety critical intervention policy is applied to the overly simplistic SIR model. In order to make predictions more reliable it is necessary to use a higher fidelity compartmental model. Moreover, doing so allows for additional safety-critical constraints to be considered, including hospitalization and death.

**Figure 2.**
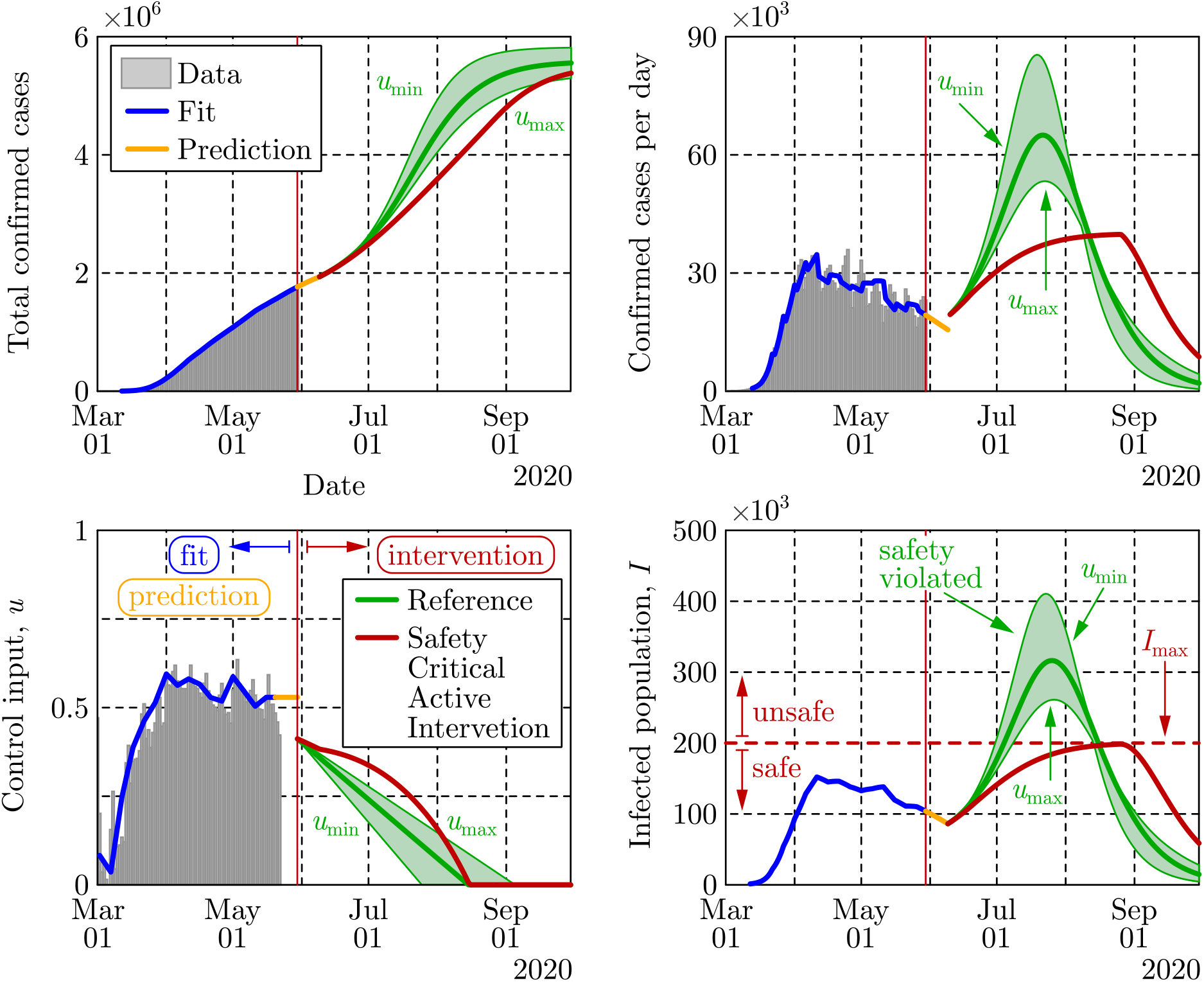
Application of the safety-critical active intervention policy in Eq. (2) that keeps the number of infected people under a given limit *I*_max_, to the SIR model in Eq. (1) with the parameters that yielded Fig. 1. The safety-critical policy is compared against a reference policy where the control input is reduced linearly. The epidemic ends relatively early due to the reduced population *N*, used in the model.

### Safety-Critical Active Interventions for the SIHRD Model

The safety-critical approach to active intervention can be applied to more complex compartmental models, viewed as control systems. To better capture other salient populations for which safety is critical, we consider the SIHRD model (detailed in Methods) which includes the *S, I* and *R* populations of the SIR model together with hospitalized and deceased populations, *H* and *D*, respectively^20, 23^. The equations governing this model are, therefore, similar to those in Eq. (1) with the addition of dynamics governing the evolution of populations associated with hospitalization and deaths. Correspondingly, the control input again appears via the time varying transmission rate *β*(*t*) = *β*_0_(1 − *u*(*t*)),while *γ* still denotes the recovery rate of the infected population. The additional parameters *λ* > 0, *ν* > 0 and *µ* > 0 represent the hospitalization rate, recovery rate in hospitals and death rate, respectively. These rates are obtained by fitting the model to the data together with the effective population *N* that becomes 13.2 million for this model (as discussed in Methods).

The evolution of the SIHRD model is shown in Fig. 3 relative to US data, including mobility data, where the fits accurately capture the data for the infected, hospitalized and deceased populations to present day (the predictive power of this model is illustrated in Methods). Also illustrated in Fig. 3 are policies that allow for future mitigation designed using the safety-critical paradigm. Safety-critical active intervention policies can be synthesized for the SIHRD model, wherein the additional compartments allow for the consideration of safety constraints aimed at limiting hospitalization and death. In particular, we will consider two active interventions policies: one policy analogous to Eq. (2) aimed at limiting the infected population, and another policy aimed at simultaneously limiting both the number of hospitalized and dead. The results of applying these two policies are shown in Fig. 3, with the specific controllers detailed in Methods. Note, additional policies could be considered, i.e., ones bounding the populations in any compartment, or any combination thereof.

**Figure 3.**
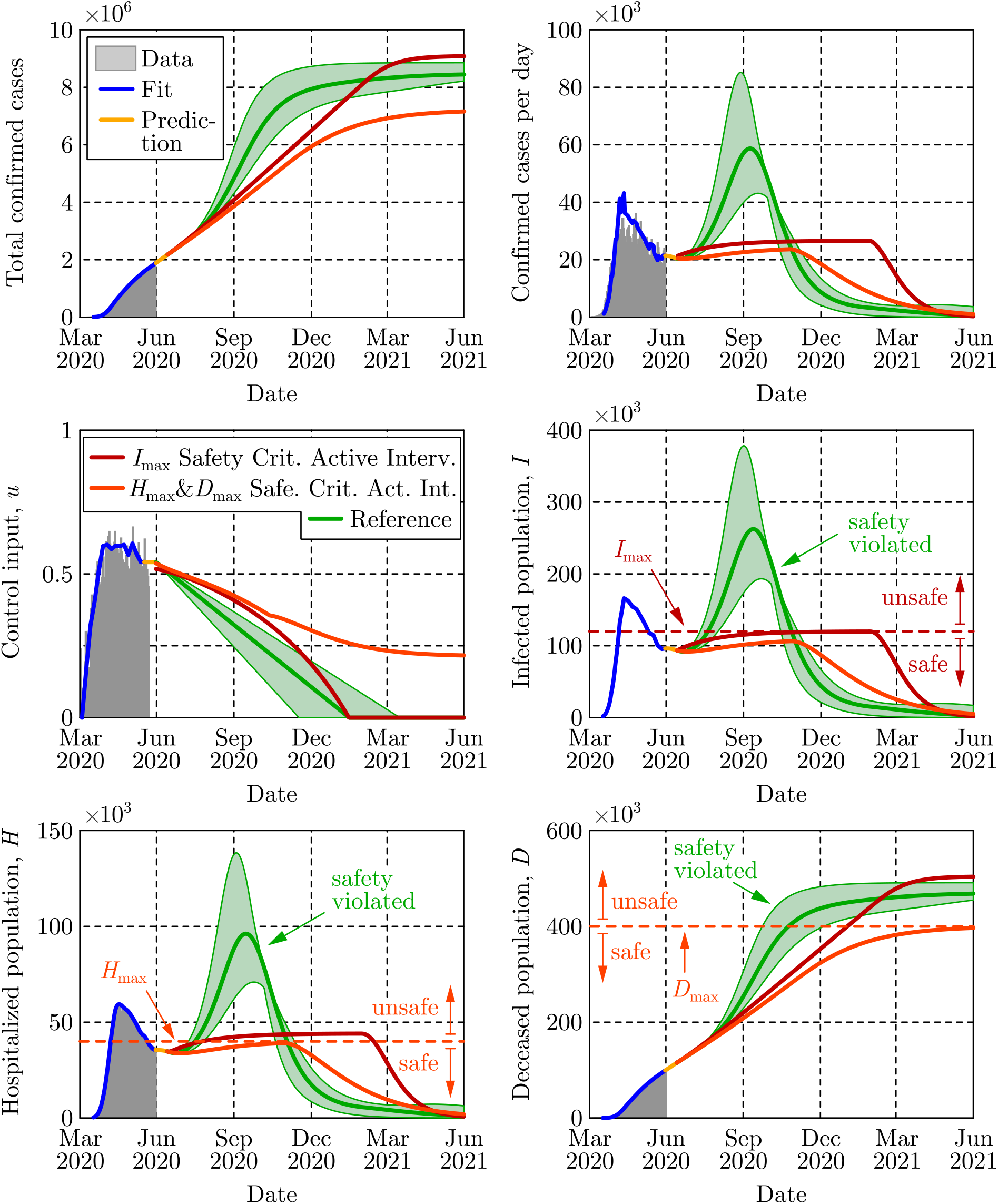
Two safety-critical active intervention policies applied to the SIHRD model. The red policy keeps the number of infected under *I*_max_ as in Fig. 2 while the dark orange policy keeps the number of hospitalized under *H*_max_ and also keeps the number of deaths under *D*_max_. The reference policy, that is linear in time, fails to maintain safety and results in a spike in infections and hospitalizations.

The first safety critical policy considered aims to limit the number of infected, i.e., *I* (*t*) ≤ *I*_max_,with results qualitatively similar to those of the SIR model in Fig. 2. Again mitigation measures are enforced over the same duration as a linear “opening up” policy while the optimality of the safety-critical policy results in substantially fewer infections at the peak. The second safety critical policy aims to limit hospitalizations (*H* (*t*) ≤ *H*_max_) based upon hospital capacity, while simultaneously limiting deaths (*D* (*t*) ≤ *D*_max_) Achieving these objectives, as indicated in Fig. 3, requires maintaining a non-zero input for a longer duration, i.e., some form of mitigation must be practiced for an extended period to limit overall death. This reflects the practices of countries that successfully mitigated the first wave of the epidemic^49^. Importantly, both of the synthesized safety-critical active intervention policies guarantee the safety constraints while simultaneously minimizing mitigation—compared against the naive linear reference policy which would drive the number of hospitalized above the limit *H*_max_, and result in large number of deceased persons. This indicates the important role that active intervention policies can play in guaranteeing safety, encoded by limiting hospitalizations and deaths.

The safety-critical policies synthesized above can also be applied to smaller geographical areas. This is especially relevant from a practical perspective, as specific mitigation efforts are determined at a state level in the US. In Fig. 4, the results are shown for four different states with safety-critical active intervention policies simultaneously bounding hospitalization and death; the safety bounds *H*_max_ and *D*_max_ were chosen as outlined in Methods, and different bounds can be used based upon state-level public policy. Different states require different levels of mitigation as highlighted by the color of each state. The gating criterion for state level mitigation was, as a proof of concept, determined by the value of the safety-critical control input 30 days after the start of active intervention; other criterion could be used based upon public policy. In this case, Michigan may open up, i.e., relax its mitigation efforts relatively quickly, reducing the control input to less than 50% of its current value in 30 days, yet mitigation efforts must be kept in place throughout the year. Qualitatively similar behavior can be seen in the case of New York, though active interventions cannot be reduced as quickly—if relaxed too quickly the result is a second spike in infections equal to the first already experienced. By comparison, California needs to very slowly relax its mitigation efforts and settle into a steady state mitigation at 80% of its current value, or the result is an outbreak with very high number of hospitalized and substantially more death. Texas should increase its current mitigation efforts to avoid a sudden and significant rise of infections, hospitalizations and death. In the case of both California and Texas, the way in which they open has a profound effect on the total hospitalizations and deaths, with deaths more than doubling if a naive opening up policy is implemented. Therefore, the safety-critical approach can determine the optimal way in which states should open—assuming good data at the state level—thereby informing policy that has the potential to dramatically reduce hospitalizations and deaths.

**Figure 4.**
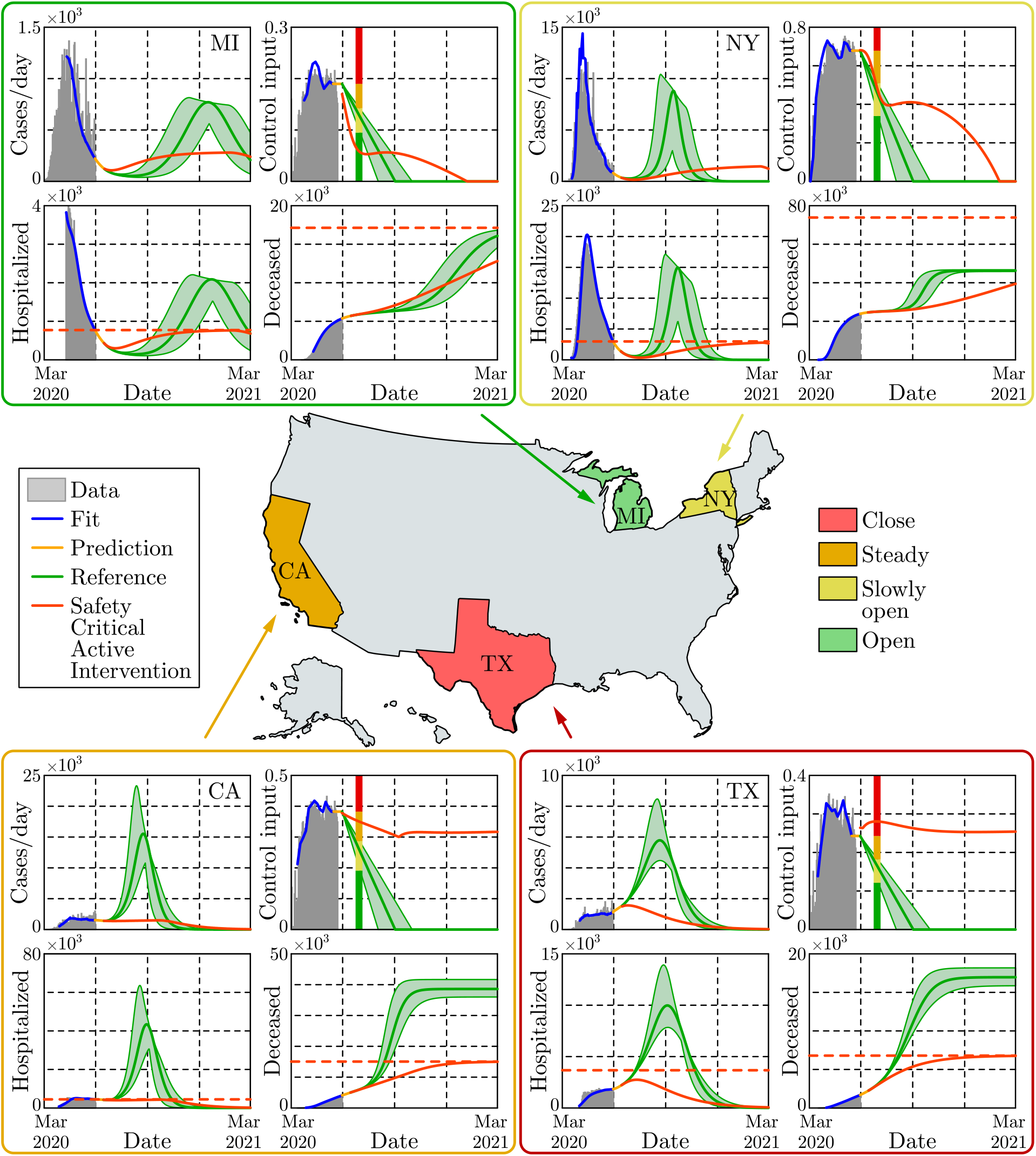
Safety-critical active interventions at the state level for four states: California, Michigan, New York, and Texas. The SIHRD model, viewed as a control system, was fit to the data for each state through May 30, 2020. From this, safety-critical active intervention policies that simultaneously bound hospitalizations and deaths are synthesized. The color of each state is determined by the control input 30 days after the start of the safety-critical active interventions, as indicated by the vertical line in the control input plots. The safety-critical policy is compared against the naive linear opening up reference policy which violates the safety bounds—resulting in over twice the deaths in the at risks states: California and Texas. This illustrates that the way in which states open up has important ramifications.

### Summary

The approach taken in this paper revolves around a new paradigm: viewing compartmental epidemiological models as control systems, viz. Eq. (1). Importantly, this perspective allows one to view these models not as systems that evolve independent of human behavior, but rather as systems where human behavior is an input that can *actively* modify their evolution (cf. Fig. 1). In this setting, we are able to synthesize active intervention policies that can serve to guide future mitigation efforts. We specifically synthesized safety-critical policies that formally guarantee that the evolution of compartmental models—the SIR and SIHRD—stay within “safe sets.” These safe sets encode bounds on the number of infected, hospitalized, and deceased populations. Closed form expressions for optimal active intervention policies were synthesized, as in Eq. (2), that ensure safety. To demonstrate this approach, US COVID-19 data on cases, hospitalizations and deaths were utilized to fit the static parameters of the SIR and SIHRD models. The active component of the control system, i.e., the control input, was synthesized utilizing mobility data; the result was models with predictive power. Projecting into the future while compensating for the incubation and testing delays, the active intervention policies were applied and compared against “naive opening up” policies. It was shown that the safety-critical policies that limit hospitalizations and deaths greatly outperformed these reference policies (Fig. 2), and this was demonstrated at both the national (Fig. 3) and the state level (Fig. 4).

### Policy Implications

The safety-critical approach to active intervention can directly inform public policy. To wit, the results presented demonstrate that epidemiological models (viewed as control systems) can capture the role of human action in mitigating COVID-19; both to describe observed data, and to actively modulate future behavior. Active intervention policies (feedback control laws) can, therefore, be used to guide non-pharmaceutical actions that should be taken to achieve a desired outcome with regard to the COVID-19 pandemic—or unforeseen future pandemics. Of particular concern are mitigation efforts devoted to ensuring safety; this encodes the desire to limit the infected, hospitalized and deceased population. The safety-critical active intervention policy presented herein results in concrete guidance on future mitigation efforts needed to achieve these guarantees. These actions can be at a local, state, national or international level depending on the ability to guide active interventions among these populations. The end result can be codified in tangible and specific public policies on “opening up”, i.e., on lifting or increasing mitigation efforts. As demonstrated throughout this paper on COVID-19 data and the corresponding epidemiological models, safety-critical active interventions—if properly encoded as public policy—have the ability to ensure available hospital capacity and save lives.

## Methods

### Safety-Critical Control for Guaranteed Safety

Safety can be framed as set invariance^50–52^ in the context of control systems and controller synthesis. Let ℛ^*n*^ be the state space of the compartmental model of interest, consisting of *n*-dimensional Euclidean space, with *n* the number of compartments, i.e., for the SIR model *n* = 3 and for the SIHRD model *n* = 5. A state x ∈ ℛ^*n*^ consists of values of the populations, e.g., x= [*S, I, R*]^T^ for the SIR model. A safety constraint is a function *h*: ℛ^*n*^ → ℛ that encodes the safe behavior of the system through:

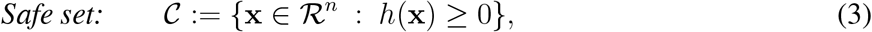

wherein the goal is for the system to evolve in this safe set. For example, for the SIR model *h*(*S, I, R*) = *I*_max_ −*I*, with the set 𝒞 containing the states for which *I* ≤*I*_max_. The goal is to give (necessary and sufficient) conditions for control systems, and synthesize corresponding policies, that render this set forward invariant, i.e., that keep the system safe.

A *control system* (in control affine form) is a first order nonlinear differential equation with a control input:

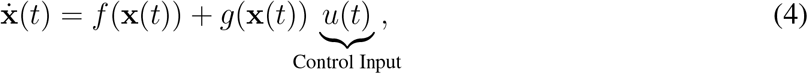

where x ∈ ℛ^*n*^ and *u* ∈ ℛ is the scalar valued control input (note that all of the methods presented also hold for vector valued control inputs). All compartmental models can be expressed in the general form of Eq. (4); which becomes an autonomous dynamical system (as they are typically modeled) for *u*(*t*) ≡0, i.e., the system evolves according to 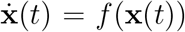. The addition of the control input *u*(*t*), as was done in Eq. (1), allows one to modify the evolution of the system to achieve desired behaviors. This modification is done via *control laws* or *policies*: *u*(*t*) = *K* (x (*t*)). The result is a *closed loop* dynamical system 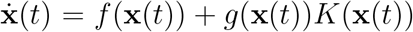, wherein x(*t*) is a solution to this system with initial condition x(0) = x_0_

We are interested in guarantees of safety framed as set invariance per Eq. (3). Thus, we say that the control system in Eq. (4) is *safe* with the policy *u* (*t*) = *K* (x (*t*)) if x_0_ ∈ 𝒞 implies that x(*t*) ∈ 𝒞 for all *t ≥*0, where x(*t*) is a solution to the closed loop system with the policy applied. By the definition of the safe set in Eq. (3), safety is thus equivalent to satisfying the safety constraint for all time: *h*(x(*t*)) ≥0. Safety-critical control addresses the fundamental question: *how does one synthesize control policies that render the set 𝒞 safe*, i.e., control policies such that safety constraint *h*(x (*t*)) ≥0 is satisfied for the closed loop system? To achieve safe behavior for the control system in Eq. (4) representing an abstract compartmental model, we leverage the framework of control barrier functions^7^. This is a new methodology for controller synthesis that has its bases in a long and rich history of set invariance for dynamical systems^50–52^. In particular, by considering the function *h*(x) that defines the safe set 𝒞, we wish to find conditions on the rate of change of this function that guarantee forward set invariance; conditions that can be checked over the entire set 𝒞 and thereby used to synthesize control policies.

It was discovered^7^ that necessary^1^ and sufficient conditions for forward set invariance are given by lower bounding the rate of change of *h* when differentiated along x(*t*) with respect to time:

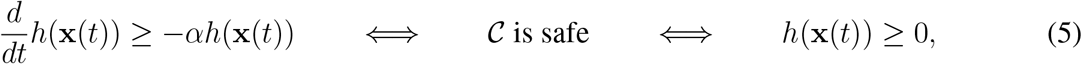

for *α* > 0 and all *t ≥* 0. The importance of the derivative condition is that it can be checked at every point of time with respect to the input *u*. Thus, *h* is a *control barrier function*^7^ (CBF) if there exits a *u*(*t*) such that:

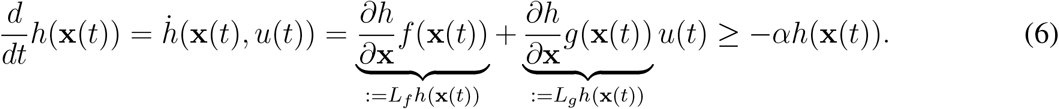

As a result, for a control barrier function, one can synthesize a policy that ensures safety by choosing a controller *u*(*t*) that satisfies Eq. (6). For example, if *L*_*g*_*h*(x) ≠ 0 then *h* is a control barrier function as *u*(*t*) satisfying Eq. (6) can be explicitly solved for through the pseudoinverse. We seek to do this in an optimal way so as to minimize the amount of active intervention.

With the goal of achieving safety while minimizing the input—as is the case with compartmental epidemiological models where we wish to minimize the active intervention—the control law synthesis problem can be framed as an optimization problem; specifically, a quadratic program:

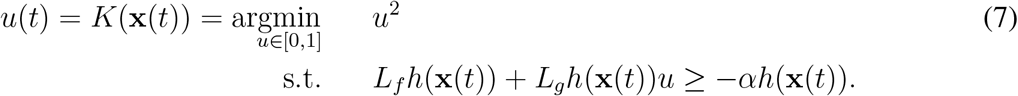

Note that here we limit *u* ∈ [0, 1] since this corresponds to the interval of active interventions with *u* = 0 denoting no intervention and *u* = 1 denoting complete intervention, e.g., fully isolating the infected population. Importantly, one can explicitly solve the optimization problem in Eq. (7) to get a closed form expression:

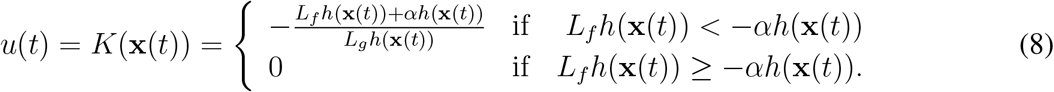

For this choice of control law, the closed loop system is safe and, additionally, the minimal input is optimally chosen. This is represented by the conditional statement, wherein *u* = 0 if the natural dynamics of the system satisfy the control barrier function condition in Eq. (6).

If *L*_*g*_*h*(x(*t*)) > 0, Eq. (8) simplifies to:

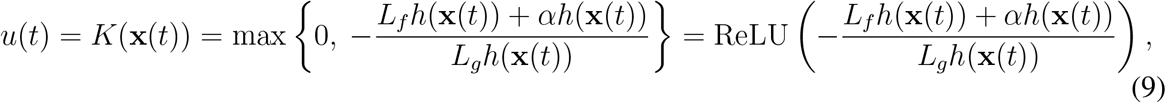

wherein the min becomes the max when *L*_*g*_*h*(x (*t*)) *<* 0. It is this formulation that leads to the active intervention policies that we will synthesize for both the SIR and SIHRD compartmental models, viewed as control systems.

### Application of Safety-Critical Methods to the SIR Model

Consider the SIR model, viewed as a control system, as given in Eq. (1). This is clearly of the form of the general control system given in Eq. (4) wherein *x* = [*S,I,R*] ^T^ ∈ ℛ^3^ and:

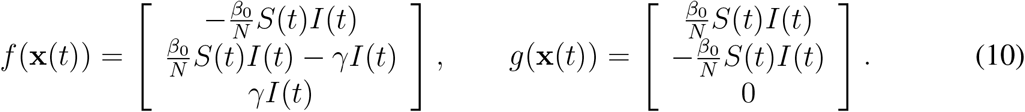

As a result, 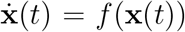 is just the standard SIR model—viewed as an autonomous dynamical system. As indicated above, the safety constraint *I* (*t*) ≤ *I*_max_ leads to the function *h* (*I*) = *I*_max_ − *I* defining the safe set 𝒞 = {[*S,I,R*]^T^ ∈ ℛ^3^: *I* ≤*I*_max_} as in Eq. (3). For the safety function *h* (*I*) = *I*_max_ − *I* calculating Eq. (6) yields:

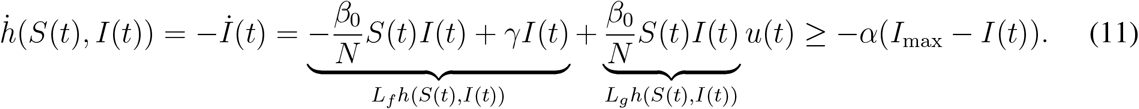

It follows that h is a control barrier function since *I* ≠ 0 and *S* ≠ 0 corresponds to having nonzero infected or susceptible populations, and therefore, *L*_*g*_*h* (*S*(*t*),*I*(*t*)) ≠ 0. The explicit solution in Eq. (9) to the optimization-based controller in Eq. (7) becomes:

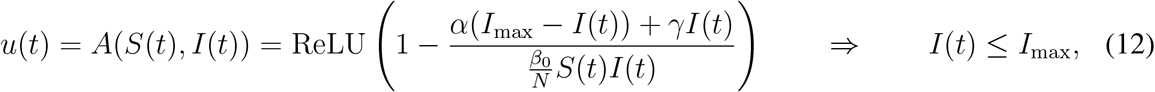

if *I* (0) ≤ *I*_max_ since in the domain of interest *S* > 0, *I* > 0 ⟹ *L*_*g*_*h*(*S*(*t*), *I*(*t*)) > 0. By picking *α* = *γ*, the control law in Eq. (12) yields Eq. (2) which was used in Fig. 2.

### Safety-Critical Control Applied to the SIHRD Model

The SIHRD model is a compartmental epidemiological model that extends the SIR model to include two additional compartments related to hospitalized and deceased populations. These additional compartments will be important in the synthesis of safety-critical controllers that bound these populations. The SIHRD model—viewed as a control system—is illustrated in Fig. 5, where *S, I* and *R* are the same populations as in the SIR model, *H* denotes the population that is currently hospitalized due to the virus (and assumed not to transmit to the susceptible population as a result), and *D* is the deceased population. The rate constants are indicated along the arrows linking the compartments: *β*_0_ is the transmission rate while *γ* and *ν* are the recovery rates of the infected and hospitalized populations, respectively. Additionally, *λ* represents the hospitalization rate and *µ* is the mortality rate. These parameters are coupled via 1*/*(*γ* + *λ* + *µ*) which is the characteristic infectious period of the virus, accounting for hospitalizations and deaths, after which there is assumed to be no transmission.

**Figure 5.**
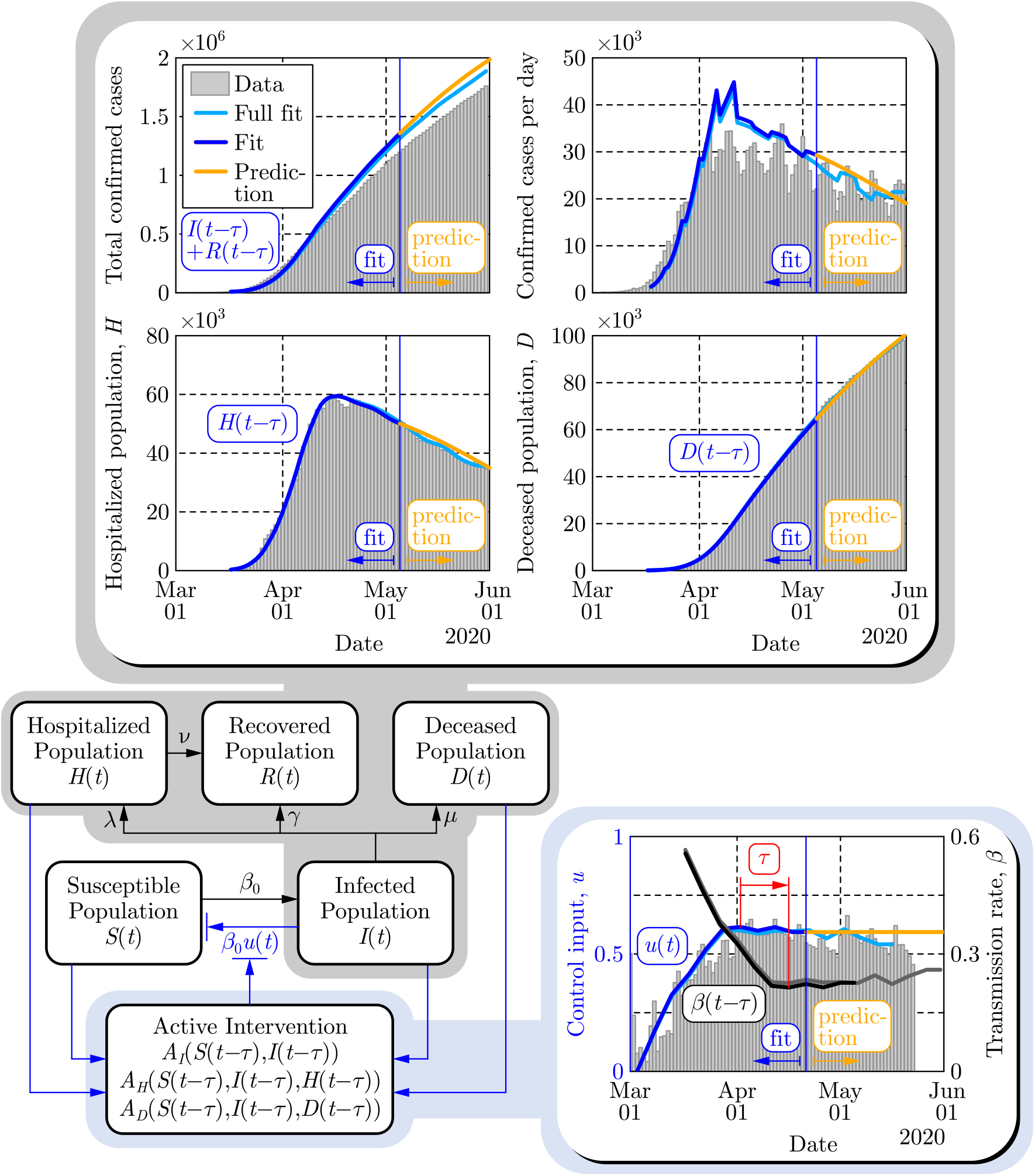
Illustration of the predictive power of the SIHRD model. The model parameters are estimated using the US data up to May 5, 2020 (dark blue), shown as a vertical blue line, and then used to predict forward for 25 days until May 30, 2020 (yellow). These are compared to a fit where the data was used until May 30 (light blue).

When casting the model in the form of Eq. (4), we use x= [*S, I, H, R, D*]^*T*^ ∈ ℛ ^5^ and obtain the following control system:

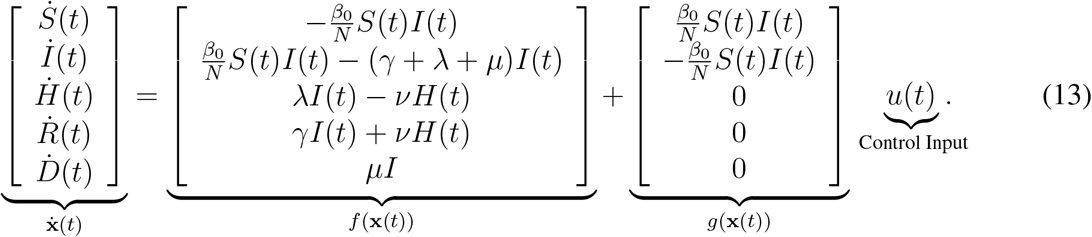

As indicated in Fig. 5, one may design active intervention policies^2^ for the control input *u*(*t*) that modulates the transmission rate: *β*(*t*) = *β*_0_(1 − *u*(*t*)). In particular, we are interested in synthesizing safety-critical active intervention policies that bound infections, hospitalization and death for the SIHRD model. The corresponding safety functions are given by:

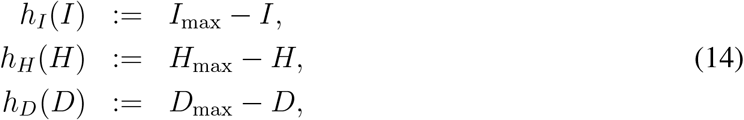

with corresponding safe sets 𝒞_*I*_, 𝒞_*H*_, and 𝒞_*D*_ defined as in Eq. (3). In the case of *h*_*I*_, a similar calculation to that in Eq. (11) yields the active intervention policy (analogous to Eq. (12)):

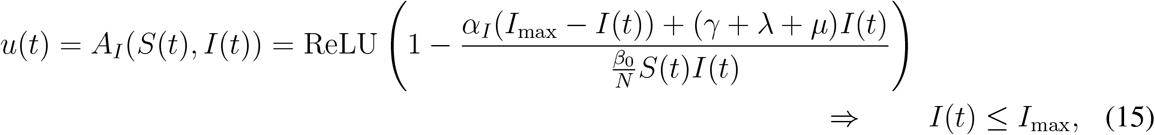

assuming *I*(0) ≤ *I*_max_, wherein we selected *α*_*I*_ = (*γ* + *λ* + *µ*)*/*10 in Fig. 3.

For the safety functions, *h*_*i*_ for *i* ∈ *{H, D}*, associated with hospitalization and death, additional steps are needed to synthesize the active intervention policy. In particular, the input *u*(*t*) does not appear when differentiating these functions as was the case in Eq. (6). Yet, we know by Eq. (5) that sufficient conditions for the sets 𝒞_*i*_ to be safe are given by 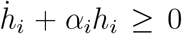 for *i* ∈ *{H, D}*, where now 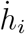 does not depend on the input *u*(*t*) as *L*_*g*_*h*_*i*_(x) = 0. As a result, define the following *extended safety functions*^50,51^:

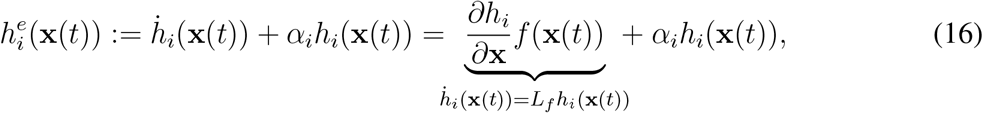

with associated safe sets: 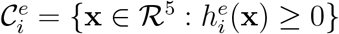. Importantly, 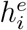 are now themselves control barrier functions, wherein the condition in Eq. (6) becomes:

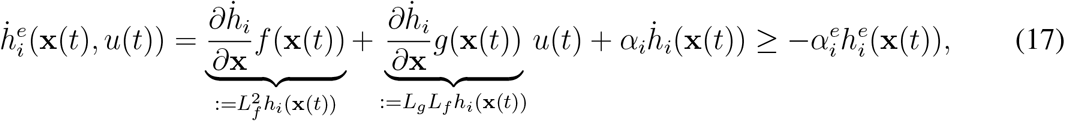

with *i* ∈ {*H,D*}

This allows us to synthesize optimal active intervention policies as in Eq. (7) via:

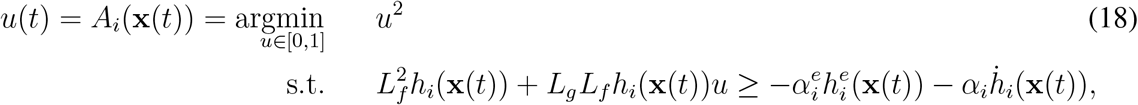

which can be converted into a conditional statement in the form of Eq. (8). Applying these constructions to the SIHRD model result in the optimal active intervention policies:

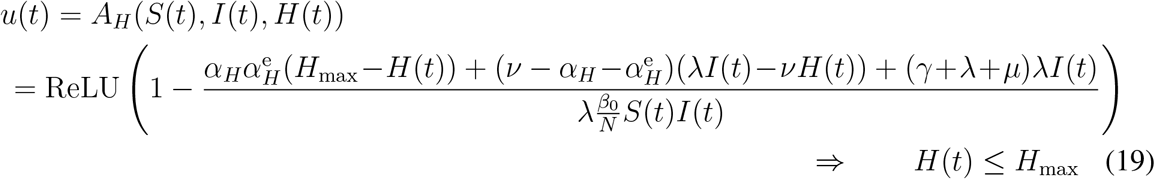

assuming the initial condition satisfies ^3^ *H* (0) ≤ *H*_max_ and 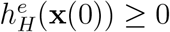, and

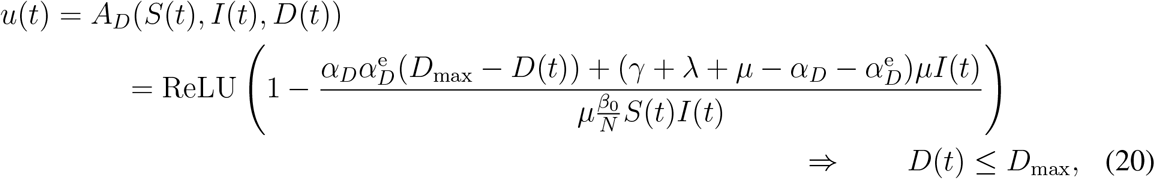

assuming *D* (0) ≤ *D*_max_ and 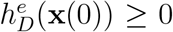. We selected 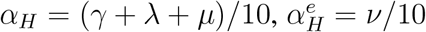 and 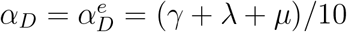 to generate Figs. 3 and 4 as described in the next section.

### Enforcing Multiple Safety Constraints Encoded as Control Barrier Functions

In addition to enforcing safety constraints via individual barrier functions, we can simultaneously enforce multiple safety constraints. We will demonstrate this in the context of enforcing both the safety constraints associated with hospitalization and death, *h*_*H*_ ≥ 0 and *h*_*D*_ ≥ 0 as given in Eq. (14), for the SIHRD model. Note the same concepts apply if we wanted to simultaneously enforce *h*_*I*_ ≥ 0 or any combination of the constraints *h*_*I*_ ≥ 0, *h*_*H*_ ≥ 0 and *h*_*D*_ ≥ 0. Similarly, these ideas can be applied to multiple safety constraints for more complex compartmental models, e.g., the SIDARTHE model^5^.

In order to limit the number of hospitalized and deceased populations in the SIHRD model, encoded by *h*_*H*_(*H*) = *H*_max_ − *H* ≥ 0 and *h*_*D*_(*D*) = *D*_max_ −*D* ≥ 0, while minimizing the active intervention *u*, we consider the quadratic program (QP):

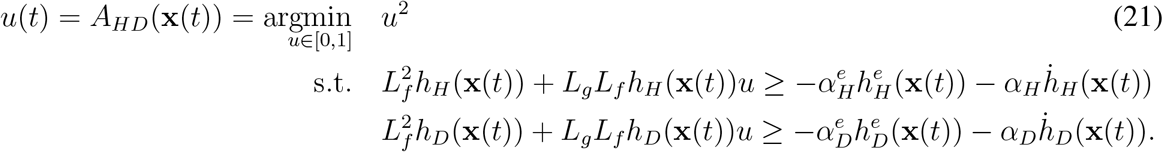

Here, we simultaneously enforce the (extended) barrier function condition in Eq. (17) for the func-tions 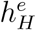 and 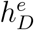 that imply satisfaction of 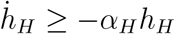 and 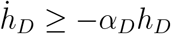 as desired. In general, it is not guaranteed that a QP with multiple constraints is feasible without a relaxation term^7^ but in this case due to the special structure of the control barrier functions considered, a solution can be guaranteed and stated in closed form.

To see this, we begin by noting that for *h*_*i*_, *i* ∈ {*I, H, D}*, defined as in Eq. (14):

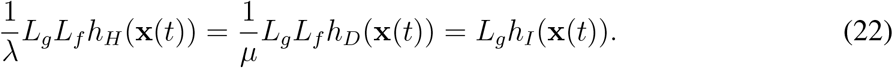

Therefore, defining

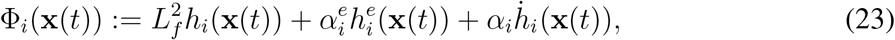

the conditions in Eq. (21) can be restated as a single inequality constraint:

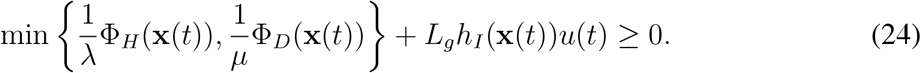

Thus, we can explicitly solve for the QP with this single constraint, yielding the same general form as Eq. (8) which leads to Eq. (9). In particular, when *L*_*g*_ *h*_*I*_ (x(t)) > 0, the result is the explicit form for a controller that simultaneously enforces *h*_*H*_ (x (*t*)) ≥ 0 and *h*_*D*_ (x (*t*)) ≥ 0 for t > 0:

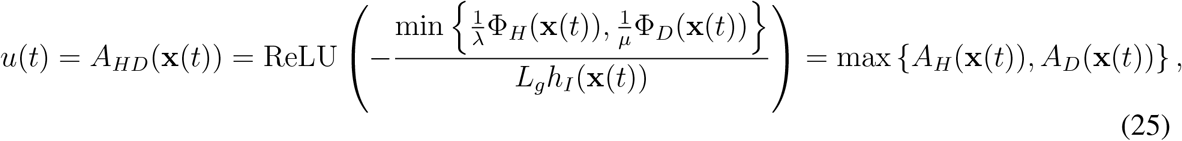

assuming *h*_*i*_ (x (0)) ≥ 0 and 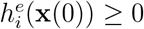 for *i* ∈ {*H, D*}Therefore, the combined conditions on bounding hospitalizations and death can be achieved via taking the maximum of the controllers *A*_*H*_ and *A*_*D*_, as given in (19) and (20), respectively. This controller is applied in Fig. 3 using US data and in Fig. 4 using state-level data (as described later in Methods). Figure 6 further demonstrates the range of safety-critical behaviors one can achieve with the active intervention policy in Eq. (25), wherein a range of maximum hospitalizations and deaths are considered leading to a range of active interventions.

**Figure 6.**
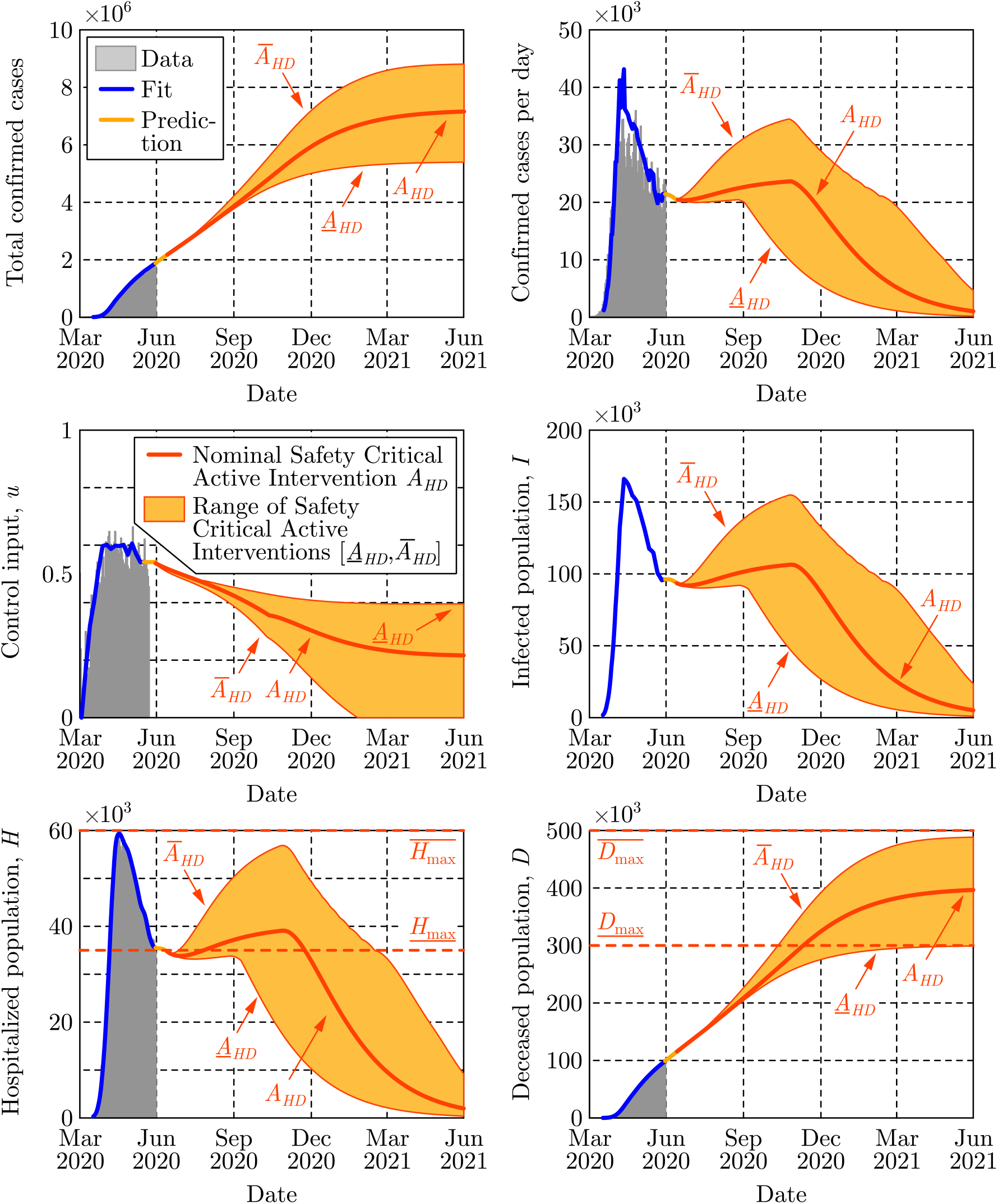
Application via the active intervention policy in E<u>q. (2</u>5) utilizing multiple barrie<u>r func</u>tions, where a range of bounds are considered for hospitalization from *<u>H</u>*<u>_max_</u> to 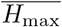, and death from *<u>D</u>*<u>_max_</u> to 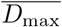. These bounds lead to safety-critical policies *<u>A</u>*_*HD*_ and *Ā*_*HD*_, respectively. These policies, and their corresponding range of interventions, are compared against the nominal safety-critical policy *A*_*HD*_ used in Fig. 3. Observe that the higher bounds on hospitalization and death, 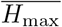 and 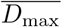, lead to the control input going to zero, while lower (and hence more stringent) bounds require sustained active intervention.

### Time Delays and Controller Synthesis with Predictors

While delays have been considered in the past in epidemic models^54–56^, they were only used to modify the autonomous dynamics of the forecasting models and thus were not considered in the context of control systems and active interventions. Here we discuss how delays affect the active intervention policies designed for the delay-free system and how to utilize predictors to compensate for the delay.

If there exists a *measurement delay τ*in a control system, cf. Eq. (4), then at time *t* only the *delayed state* x (*t − τ*) is available via measurements, while the *instantaneous state* x(*t*) of the system is unknown. Therefore, the controller must rely on the delayed state, which modifies the control law from *u*(*t*) = *K*(x (*t*)) to *u*(*t*) = *K*(x (*t* − *τ*)), yielding the time-delayed closed loop system:

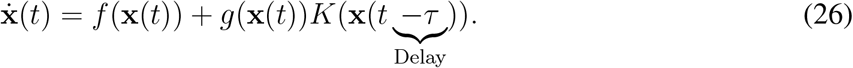

Therefore, since measurement delay affects the dynamics of the closed loop system via time delays, they are typically undesirable—especially since they are often source of instability or reduced performance^57^. In the context of safety-critical control, as considered herein, time delays may lead to violation of the safety condition if one designs a controller by assuming no delays. If the delay is large, the measured delayed state may be significantly different from the instantaneous state due to the evolution of the system over the delay interval. This prevents the delay-free control design from guaranteeing safety.

A possible solution to overcome the poor performance caused by delays is the application of *predictor feedback control* ^48, 58–61^. Predictor feedback utilizes an *internal model* in order to predict the current instantaneous state of the system from delayed measurements. Prediction is made over the delay interval based on the delayed state x(*t* − *τ*) resulting in a *predicted state* x_p_(*t*) that is an estimation of the instantaneous state x(*t*), i.e., x_p_(*t*) ≈ x(*t*). Then, one can utilize the predicted state in the controller designed for the delay-free system. This leads to *u*(*t*) = *K*(x_p_(*t*)) yielding:

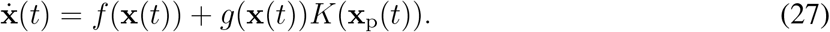

If the prediction is accurate, i.e., x_p_(*t*) = x(*t*), it eliminates the delays from the system.

To provide the predicted state x_p_(*t*) to the controller, one needs to anticipate how the closed loop system evolves under the predictor feedback controller over the delay interval [*t* − *τ, t*]. This can be achieved by introducing *θ* ∈ [*t* − *τ, t*] and substituting *t* = *θ* and x(*θ*) = x_p_(*θ*) into Eq. (27):

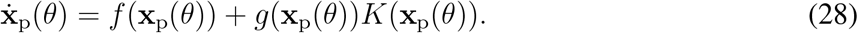

This equation is the *internal model* used for prediction. To obtain the predicted state x_p_(*t*) required at time *t* by the controller *u*(*t*) = *K*(x_p_(*t*)), Eq. (28) can be numerically integrated over *θ* ∈ [*t* − *τ, t*] with initial condition x_p_(*t* − *τ*) = x(*t* − *τ*) consisting of the most recent available measurement. Assuming a reasonable model, x(*θ*) ≈ x_p_(*θ*) for all *θ* ∈ [*t* − *τ, t*] wherein x(*t*) ≈ x_p_(*t*), and so the predictor eliminates the time delay from the closed loop system in Eq. (27).

Figure 2 was generated by directly utilizing Eq. (28) as internal model. Prediction can be further improved by noticing that during the initial time *θ* ∈ [*t* −*τ, t*_0_] (if *t* ∈ [*t*_0_, *t*_0_ + *τ*]), the system is not yet affected by an active intervention starting at *t*_0_. Thus, one can apply the nominal (fitted) model with the corresponding fitted control input (see the yellow curve in Fig. 2) to calculate the predicted state during this initial interval, and Eq. (28) can be used afterwards. In Figs. 3 and 4, this more accurate prediction algorithm was utilized, although using Eq. (28) only could already compensate the effects of the time delay and managed to maintain safety in Fig. 2.

### Predictors for Compartmental Models

In the control of COVID-19, the measurement delay originates from the incubation period and testing, and it can go up to about two weeks (*τ* ≈14). Over these two weeks, the number of infected population may have increased significantly. As a result, the infected, hospitalized and deceased populations may be much closer to the safety limit than what the data shows. Thus, active intervention policies should be applied earlier than suggested by the delay-free controller, otherwise the populations of interest may overshoot and increase above the safe limit. The predictor feedback control technique accounts for the two-weeks measurement delay by predicting what could be the true number of infected population currently. Then, the delay-free control law can be applied utilizing the predictor, and it will therefore maintain safety if the prediction is accurate enough.

Accurate prediction requires knowing the delay. As shown in Fig. 1, the delay can be identified by model fitting to compartmental data and by utilizing mobility data. Furthermore, prediction requires an accurate internal model. For the SIR model, one can utilize the closed loop system:

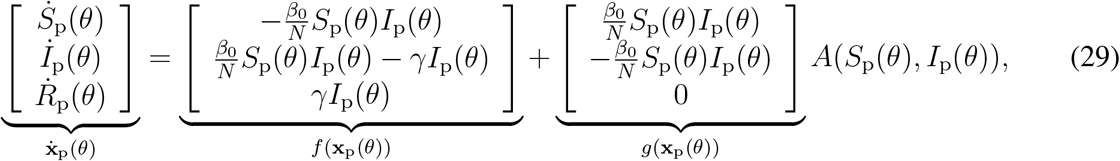

which can be integrated over the interval *θ* ∈ [*t* −*τ, t*] with initial conditions *S*_p_(*t* − *τ*) = *S*(*t* − *τ*), *I*_p_(*t* − *τ*) = *I*(*t − τ*) and *R*_p_(*t* − *τ*) = *R*(*t* − *τ*) to get the predicted states *S*_p_(*t*) and *I*_p_(*t*) required by the controller at time *t*. Similarly, for the SIHRD model prediction can be made using:

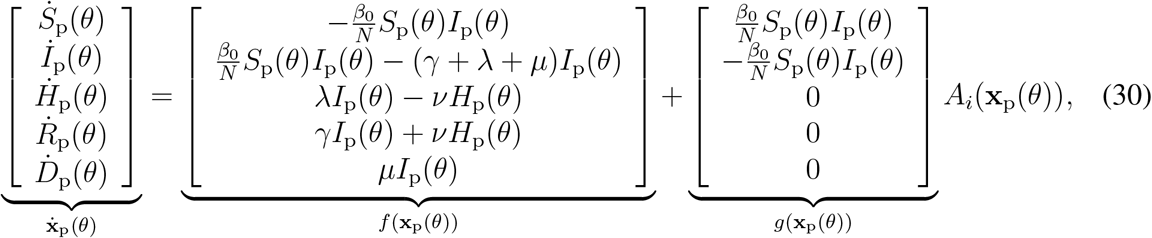

for *i* ∈*{I, H, D, HD}* depending on which active intervention policy is being utilized, i.e., whether we wish to bound infections, hospitalizations, deaths, or a combination thereof. According to Fig. 5, a short term (i.e., two-week) prediction can be made accurately with such model. Therefore, the result is safety of the system even in the presence of the time delay.

### Parameter Identification for Compartmental Models

As mentioned previously, a wide variety of model parameters can be used to fit COVID-19 data, yielding drastically different predictions for the evolution of the different populations. In order to provide a model with high predictive ability, the parameters must be determined in a way that reflects their physical meaning while the model reproduces the available data. Since the model is being treated as a control system, the sequence of inputs that reflect the intervention applied thus far can be estimated. To ensure that the estimated inputs are realistic, they are initialized using mobility data from SafeGraph^38^, a data company that aggregates anonymized location data from numerous applications in order to provide insights about physical places. Such data allow us to quantify the increased time people stay at home to increase social distancing. To enhance privacy, SafeGraph excludes census block group information if fewer than five devices visited an establishment in a month from a given census block group.

The data fitting problem identifying the parameters of the SIR model in Eq. (1) can be formulated as an unconstrained, nonlinear optimization problem:

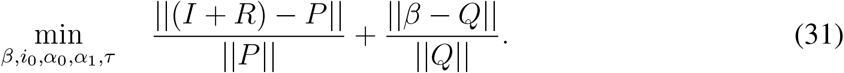

The first term in the objective function seeks to minimize the difference between the total cases (*I* + *R*) given by the model and the number of positive test cases from the data *P*. The second term seeks to minimize the difference between the transmission rate *β* and some function *Q* that models the expected transmission rate based on mobility data. To obtain (*I* +*R*) at different time moments, we integrate Eq. (1) and this integration results in the nonlinearity in the objective function.

The optimization problem in Eq. (31) is used to find the parameters of the SIR model in Eq. (1) as follows. The first decision variable is the vector *β* ∈ ℝ^*k*^, *k* = floor(*T/K*), representing the time-varying transmission rate *β*_*j*_ = *β*_0_(1 −*u*(*t*_*j*_)), *j* ∈ {1, 2, …, *k*}. This corresponds to a control input *u*(*t*_*j*_) that is updated every *K* days over the time span *T*. The value *K* = 5 days was used in order to prevent overfitting and account for the inability to define new policies on a daily basis. The decision variable 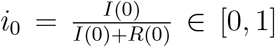 signifies the initial values of different populations, while decision variables *α*_0_, *α*_1_, *τ*are used to scale and delay the mobility data according to:

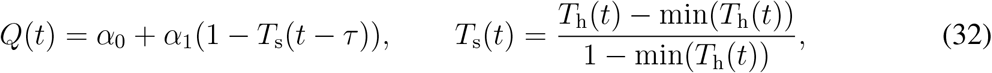

where *T*_h_ is the median percentage time spent at home. Recall that the delay *τ*exists due to the testing delay and the incubation period of the virus^42^, and it is bounded to be between 5 to 16 days. Finally, the parameter *β*_0_ is extracted from *β* and *Q* as *β*_0_= max_*j*∈{1,…,*k*},*t*∈[0,*T*]_ {*β*_*j*_,*Q* (*t*)} When fitting the SIR model to the US national data we obtain *β*_0_ = 0.51 1/day and *τ* = 10 days, while we set the parameter *γ* = 0.2 1/day to correspond to the average characteristic recovery time observed in the data. The fit for the SIR model is shown in Fig. 1 where the prediction along the 10 day period of the delay *τ* is highlighted. These predictions are used in Fig. 2 when applying the safety-critical active intervention policy in Eq. (2).

It is important to note that for the SIR model, and the corresponding optimization problem in Eq. (31), *N* is fixed at a value below the total population. This is necessary due to the underreporting of infections^43^, along with the fact that many people will never have any contact with infected individuals. Additionally, setting *N* to be the entire population of the US in the SIR model would require many years for the virus to die out even with strict social distancing. Since our goal with the SIR model was to demonstrate the concepts of safety-critical active interventions, rather than to predict the size of the populations accurately, we decided to fix *N* to be 7.5 million. Alternatively, one can make *N* a decision variable—this did not improve results for the SIR model, but will prove useful for the higher-fidelity SIHRD model as explained below.

Analogous to the optimization problem for the SIR model, the optimization problem to estimate the parameters of the SIHRD model in Eq. (13) can be framed as:

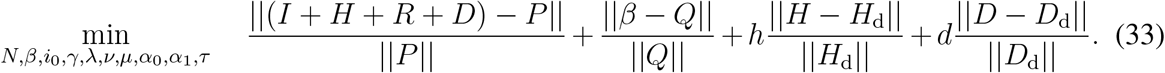

This formula accounts for the number of hospitalizations *H*_d_ and the number of deaths *D*_d_ in the data. The weights *h* > 1 and *d* > 1 are set to reflect that the data for hospitalizations and deaths are inherently less uncertain than the data on total positive test cases. In order to obtain the populations *I* + *H* + *R* + *D, H*, and *D* as function of time we integrate the model in Eq. (13) which contains the new decision variables *N, γ, λ, ν, µ*. Due to the increased complexity of the model, the parameter *γ* is no longer fixed, as it was in the SIR model, instead, the value of 1*/*(*γ* + *λ* + *µ*) is implicitly constrained to be roughly within 2.5-7.5 days through individual bounds on the decision variables. Lastly, the characteristic time at which someone recovers from the hospital 1*/ν* is bounded between 2.5 and 4 days. Finally, differing from the SIR model optimization, the population *N* is now a decision variable with a lower bound of 4% and an upper bound of 10% of the total population of the nation or state of interest. We note that the value of *N* does not change the qualitative conclusions regarding safety-critical active interventions.

These optimization problems were solved using the pagmo^62^ C++ library, and the solutions were verified using a variety of its solvers including CMA-ES, differential evolution, NSGA-II, and several solvers in the NLOPT suite. Table 2 provides the obtained parameters as computed by CMA-ES with a population size of 400 and a generation number of 400. The predictive power of the SIHRD model is illustrated by the fit in Fig. 5 over a 25 day horizon which provides accurate predictions for both the hospitalized *H* and the deceased *D* populations. This tight fit on *H* and *D* comes at the cost of a worse fit on the infected populace *I*, as reflected in the graph on total confirmed cases. However, as noted previously, the uncertainty in the total case number data is much higher than that of the *H* and *D* compartments, so this mismatch is not surprising. In fact, it could be argued that the actual number of infected persons is higher than reported and this model, as a result of its utilization of hospitalizations and deaths, actually captures this higher number. For example, in Fig. 5 the SIHRD model predicts a higher peak in the number of cases per day in the first half of April—this better describes the hospitalization and death data, and could provide a more accurate picture of cases per day due to the lack of testing at that time.

**Table 1.** Policy summary.

| Background | Main Findings | Policy Implications |
| --- | --- | --- |
| Epidemiological models provide a powerful tool to guide the mitigation of COVID-19. Yet these models are dynamical systems that must be constantly updated as new data is assimilated. Policy decisions, therefore, fail to account for future active interventions, e.g., social distancing and stay-at-home orders, that can change the evolution of these models. | Viewing epidemiological models as control systems allows for the design of active intervention policies that can mitigate COVID-19 while modulating these mitigation efforts as function of time. With this viewpoint, safety-critical policies are synthesized that guarantee safety of the system by bounding the infected, hospitalized, and deceased populations. | The safety-critical active interventions synthesized can be used to formulate and inform public and governmental policy on lifting or increasing mitigation efforts in a centralized and decentralized fashion. These policies may, therefore, guide mitigation efforts at local or national levels to ensure hospitals do not reach capacity, and overall deaths are limited. |

**Table 2.**
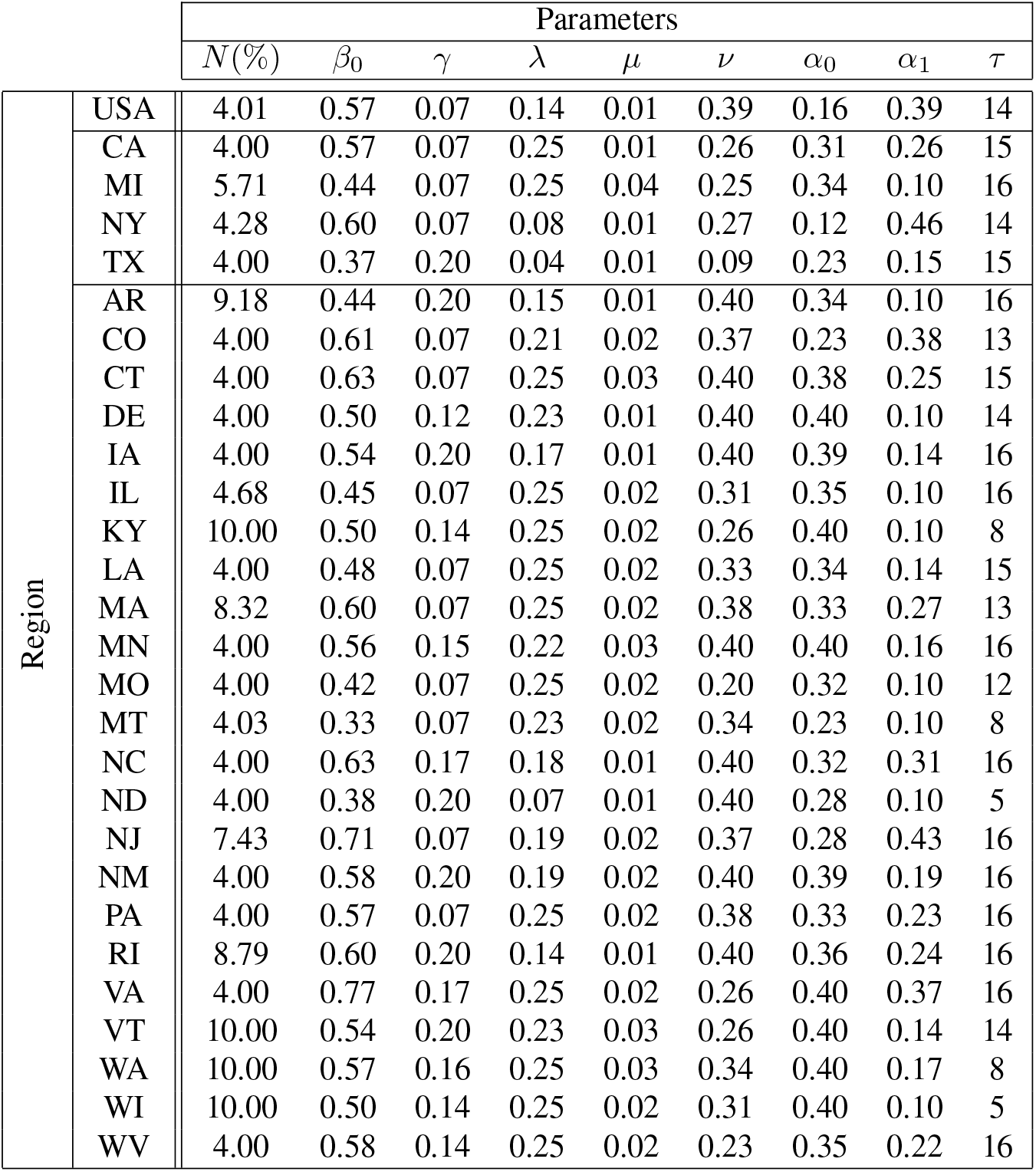
Parameter estimates for the SIHRD model in different states, including the national level, obtained by solving the optimization problem in Eq. (33). Rough agreement between the states can be seen, but parameters vary due to differing exposure levels, social distancing protocols, population dynamics, and testing capacity. Note that the five states with a significantly smaller *τ* have values shifted by roughly one week from the average, due to the cyclical nature of the data.

### State-Level Safety-Critical Active Interventions

To provide a case study in using the safety-critical control framework to determine public policy, we apply the approach described in the main body of the paper, and detailed in Methods, to all states of the US that have sufficient data. A state is considered to have sufficient data if it has recorded values of positive cases, hospitalizations, and deaths for 50 consecutive days. While the model parameters can be estimated over shorter time periods, a sufficient window of data to verify the prediction accuracy was also required. To obtain the SIHRD model parameters at the state level, the optimization problem in Eq. (33) was solved using the CMA-ES algorithm with *h* = 2 and *d* = 4 to reflect the relative certainty of the data on hospitalizations and deaths, and thus, the desire to fit the former two populations more accurately. The parameters are identified per the methods described above, with the values shown in Table 2. Utilizing the fitted models, we develop a proof-of-concept reopening policy.

To develop a template policy on whether a state should reopen, and if so how aggressively, we apply the safety-critical framework to the SIHRD model for each state. Specifically, we utilize the safety-critical active intervention policy *u*(*t*) = *A*_*HD*_(x (*t*)) given in Eq. (25). In this policy, there are two constants to be chosen: the maximum number of hospitalizations *H*_max_, and the maximum number of deceased *D*_max_. The value of *H*_max_ for each state was chosen to be the last fitted hospitalization value (i.e., the last point of the blue curves in Figs. 4 and 9) to prevent hospitalizations from getting worse than the most recent recorded data. To guarantee safety, per Eq. (19), the initial condition 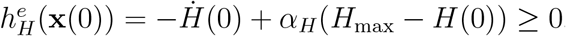. If *H*_max_ did not satisfy this (which is the case for states where the most recent value of hospitalizations is the largest), *H*_max_ is increased to the smallest feasible value: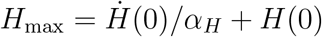 to ensure the feasibility of safety-critical active intervention. The value of *D*_max_ was chosen to be three times the value at the start time of the active intervention (i.e., the start point of dark orange curves in Figs. 4 and 10). Although this choice is arbitrary, it satisfies the condition 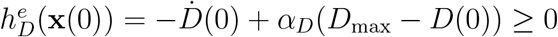 for all states and allows us to keep consistency throughout the states.

The proof-of-concept reopening framework is based upon the behavior of the control input *u*(*t*). Since this safety-critical policy is (pointwise) optimal, i.e., solves the optimization problem in Eq. (21) at each *t*, there is no better instantaneous mitigation approach. Therefore, if this policy says to increase mitigations then a state should close down and if this policy rapidly reduces interventions the state can continue to open. To translate the control input into an applicable policy we utilize a “gating criterion” that is based on the value of the control input 30 days after the beginning of the active intervention period; see the colored vertical lines in Figs. 4 and 7 that determine the colors of states on the map. If the value of the control input after 30 days is greater than its initial value, the state should close down (states in red). If the input has reduced by less than 25%, mitigation efforts should be kept steady (states in orange). If the control input has decreased by more than 25% but less than 50%, the state can slowly reopen (states in yellow). Finally, a state where the input has decreased more than 50% in this 30 day period can reopen at a more rapid rate (states in green). While this is a very simplistic policy, created to provide uniformity across states, it demonstrates one way to utilize safety-critical methods to inform public policy. Indeed, using the entirety of the control input can result in more informed state level policy decisions.

**Figure 7.**
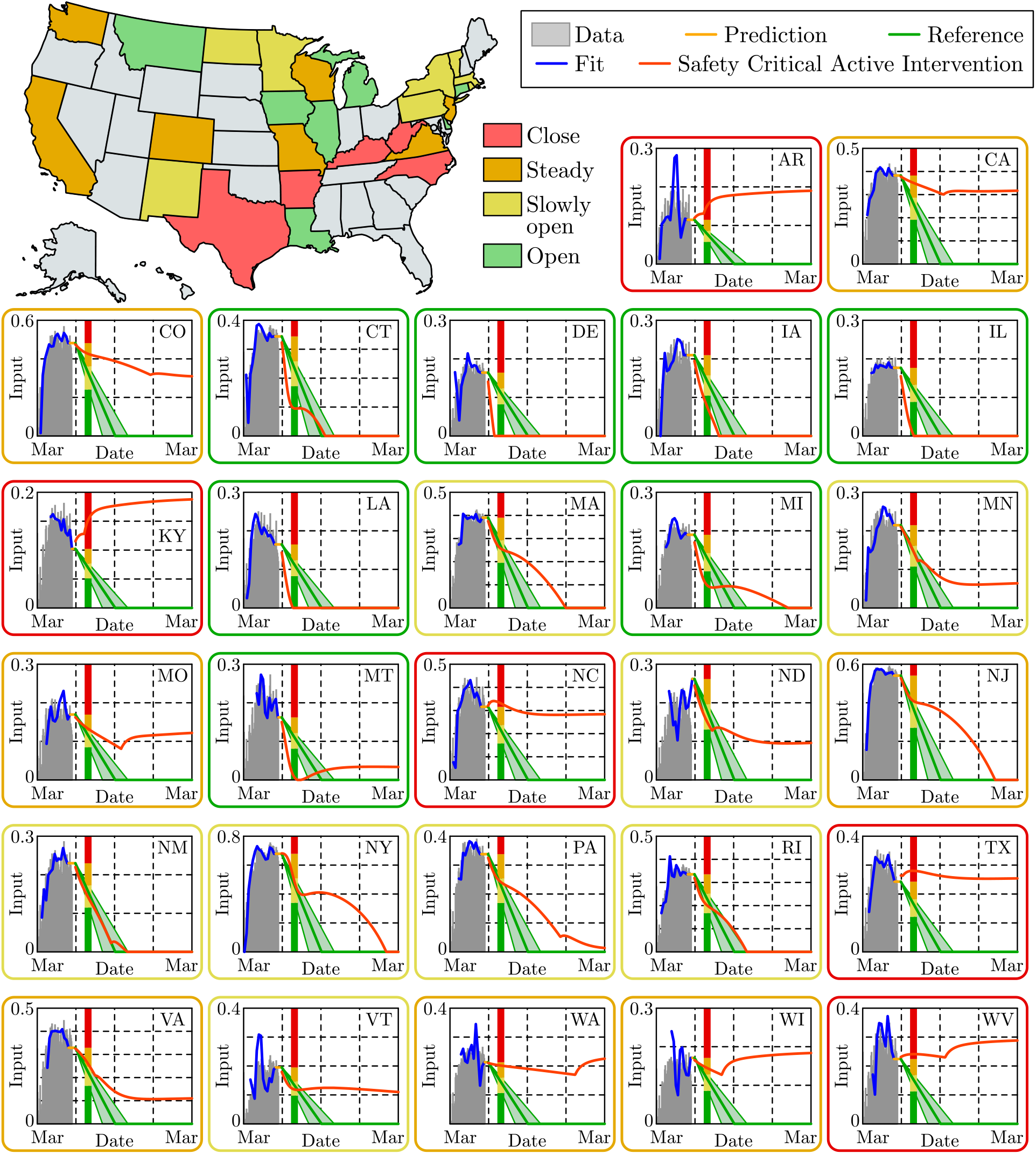
State level active intervention policies, along with the control input fit to US mobility data (grey) through May 30, 2020. Based upon the optimal safety-critical control policy (dark orange), state level mitigation efforts are recommended, i.e., a 30 day gating criterion is utilized where if at this time the safety-critical policy is increasing the state should close (red), if it has decreased by less than 25% it should hold mitigation efforts steady (orange), by less than 50% it should slowly reopen (yellow), and more than 50% it can open at a faster pace (green). This is compared against the naive linear reopening policy serving as a reference (green tube); this reference will be used in subsequent figures to demonstrate the consequences of a naive opening.

The criterion outlined above was applied to state level SIHRD models, with the results shown in Figs. 7-10. The safety-critical control inputs, that determine the state by state recommendations, are shown in Fig. 7. Also shown in that figure is the reference naive reopening policy that decays linearly in time, reaching zero on September 1 (the start of the school year). Fig. 8 shows the cases per day for the safety-critical and reference intervention policies; a second spike in cases can be seen for states that are yellow, orange and red for the reference policy. This spike is also seen in Fig. 9 in the context of hospitalizations, where *H*_max_ is exceeded for the reference policy for all states while the safety-critical policy does not exceed *H*_max_ as the theory implies. Finally, Fig. 10 shows the total deaths, wherein large death rates are seen—especially in states in red— for the reference policy while the safety-critical policy does not exceed *D*_max_. These figures, therefore, show that safety-critical control of active intervention can be used to synthesize state level reopening policies and, if followed, can limit hospitalizations and deaths.

**Figure 8.**
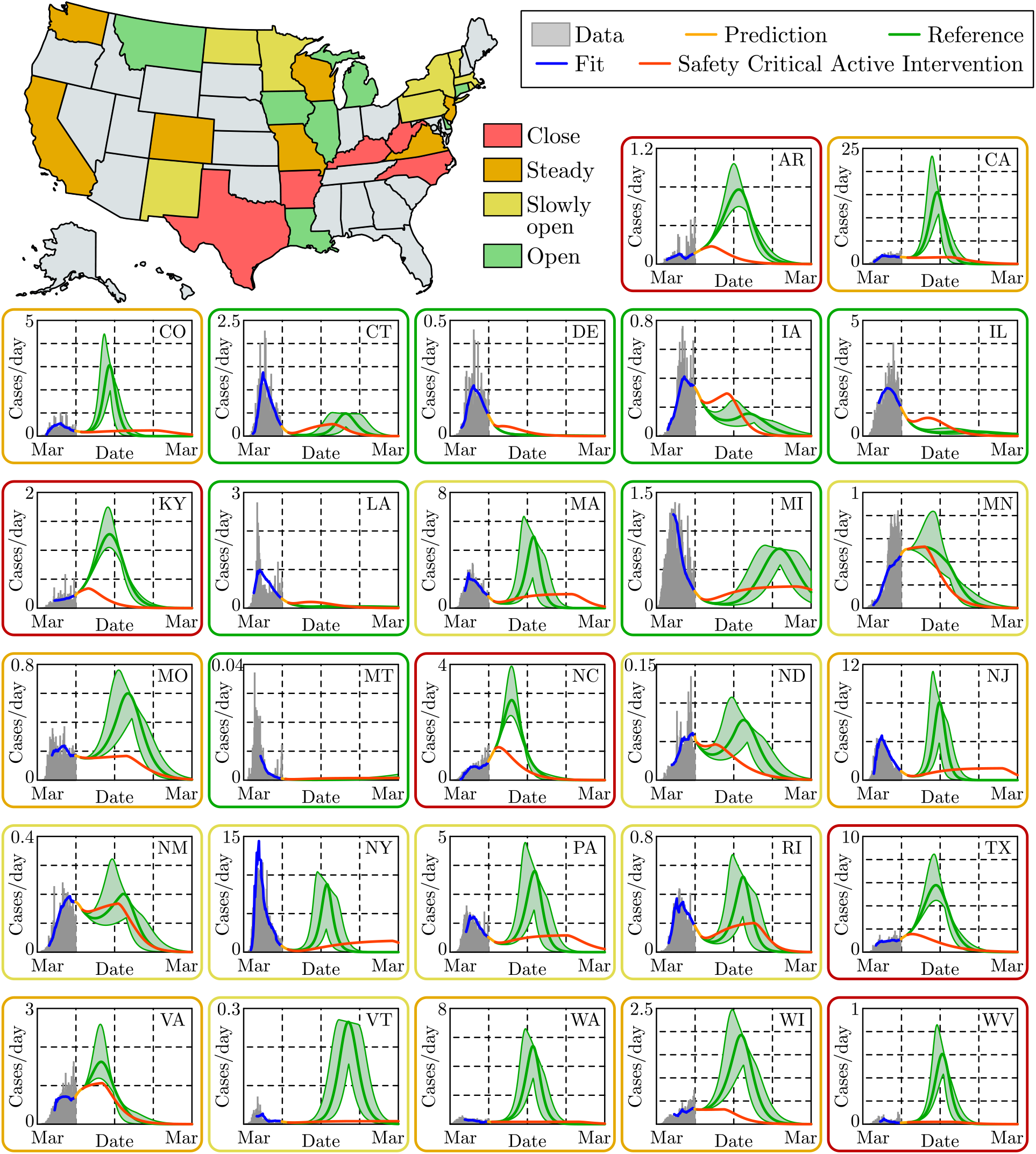
State level prediction for the cases per day over a year time span. The vertical axis measured in 1000’s of people, and state level data (grey) is shown through May 30, 2020 along with the fit from the SIHRD model (blue). The effects of different active intervention policies are shown: the optimal safety-critical active intervention policy (dark orange) and the linear naive reopening policy (green). Both of these policies, at the state level, are shown in Fig. 7. In almost all states, the result is a second spike in cases—often larger than the original spike—if a naive reopening policy is used.

**Figure 9.**
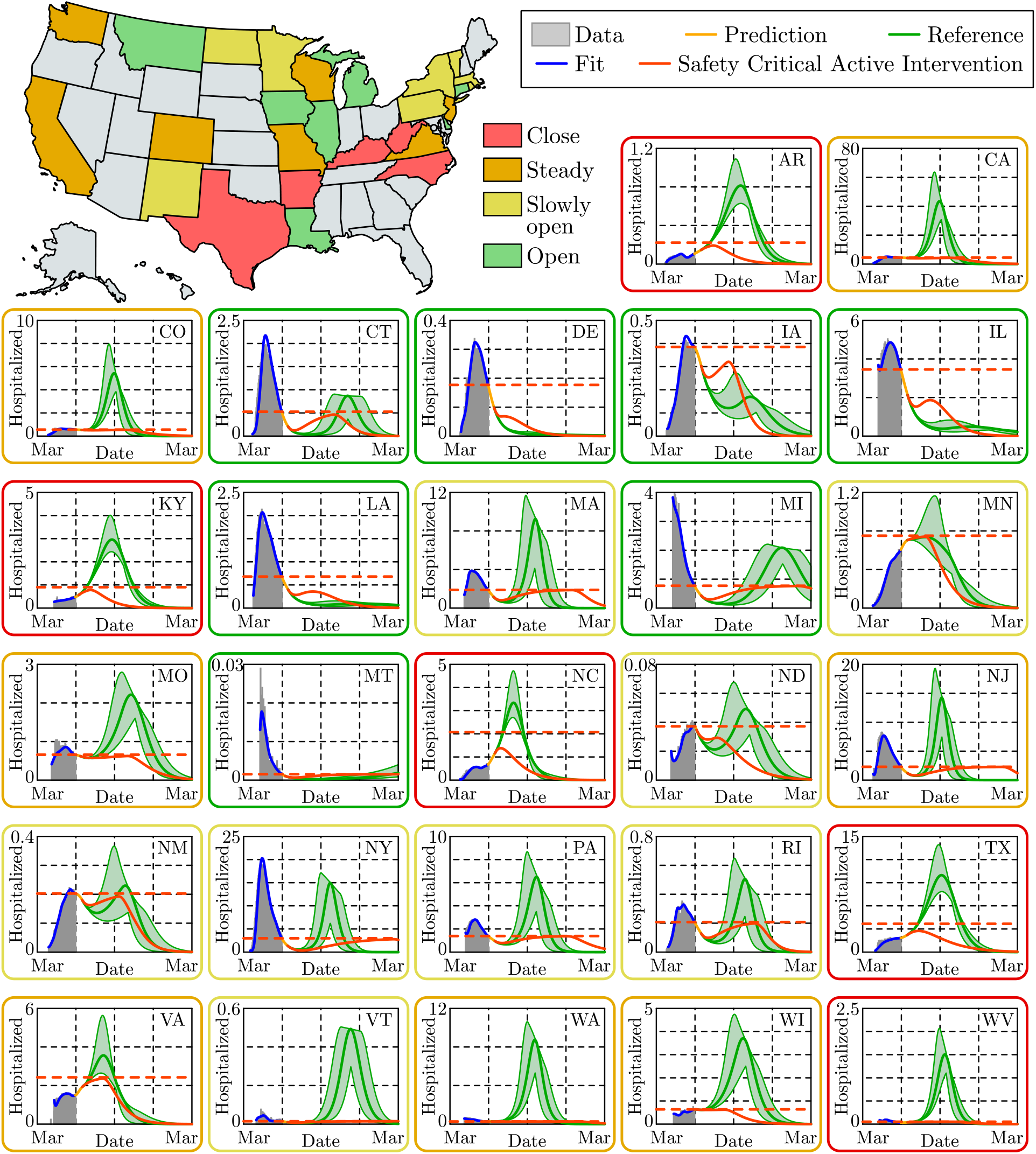
State level predictions for the number of persons hospitalized, with the vertical axis measured in 1000’s of people. State level data (grey), along with the corresponding fit of the SIHRD model (blue) for hospitalizations; note the accurate fit of this data. The optimal safety-critical active intervention (dark orange) is compared against the naive reopening reference policy (green), with both policies shown in Fig. 7. Of particular note, for the safety-critical policy the number of hospitalizations is guaranteed to be bounded above by *H*_max_ (horizontal dashed dark orange line), and it can be seen that this safety constraint is satisfied for all states. This can be compared against the reference policy where *H*_max_ is exceeded for all states colored yellow, orange and red and even some states colored green. This indicates that without careful mitigation policies, hospital capacity constraints can be easily violated.

**Figure 10.**
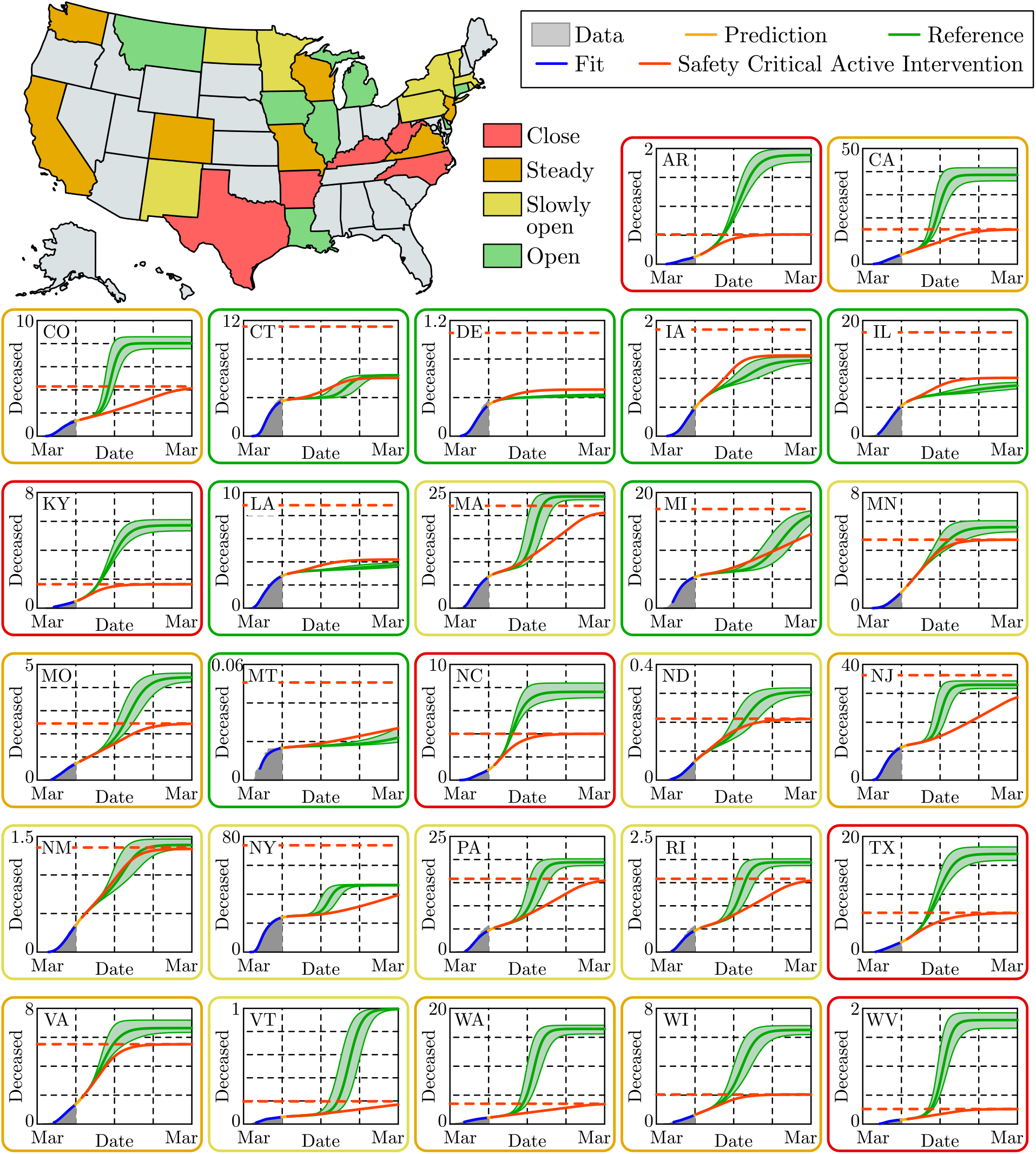
State level predictions for the total number of deaths, with the vertical axis measured in 1000’s of persons. The data for the number of deceased persons as recorded through May 30, 2020 (grey) is shown along with the fit of the SIHRD model (blue); as with the hospitalization data in Fig. 9, the fit of the data is remarkable. The results of applying the safety-critical active intervention policy (dark orange) and the naive reference policy (green), where these policies are shown for each state in Fig. 7. The safety-critical policy guarantees that the total number of deaths stays under the upper bound *D*_max_, indicated by a horizontal dashed dark orange line. Note that for states that should close down as determined by the the algorithm illustrated in Fig. 7, i.e., the states indicated in red, there is a dramatic difference in the total number of deaths between the safety-critical and reference policies indicating the essential role of proper mitigation.

## Data Availability

The data on cases, hospitalizations, and deaths comes from the COVID Tracking project.
The data on social distancing comes from SafeGraph

https://www.safegraph.com/

https://covidtracking.com/

## Acknowledgements

The authors would like to thank Franca Hoffmann for her insights into compartmental epidemiological models and Gábor Stépán for discussions regarding non-pharmaceutical interventions in Europe. This research is supported in part by the National Science Foundation, CPS Award #1932091.

## Footnotes

1 Technically, for necessity, *α* must be chosen to be an extended class 𝒦 function^7^ not a constant. We utilize a *α* constant for simplicity of exposition and without loss of generality.

2 Note that the delayed values of the different populations appear in the corresponding feedback laws which will be compensated using predictors based on the fitted model. This will be described in detail later in Methods.

3 For the extended control barrier functions ^50,53^ x (0) ∈ 𝒞 ⋂ 𝒞 ^e^ implies that x (0) ∈ 𝒞 ⋂ 𝒞 ^e^ for all *t* ≥ 0.

